# Decoding biological age from face photographs using deep learning

**DOI:** 10.1101/2023.09.12.23295132

**Authors:** Osbert Zalay, Dennis Bontempi, Danielle S Bitterman, Nicolai Birkbak, Derek Shyr, Fridolin Haugg, Jack M Qian, Hannah Roberts, Subha Perni, Vasco Prudente, Suraj Pai, Andre Dekker, Benjamin Haibe-Kains, Christian Guthier, Tracy Balboni, Laura Warren, Monica Krishan, Benjamin H Kann, Charles Swanton, Dirk De Ruysscher, Raymond H Mak, Hugo JWL Aerts

**Affiliations:** Artificial Intelligence in Medicine (AIM) Program, Mass General Brigham, Harvard Medical School, Boston, United States of America; Department of Radiation Oncology, Brigham and Women’s Hospital, Dana-Farber Cancer Institute, Harvard Medical School, Boston, United States of America; Division of Radiation Oncology, Queen’s University, Kingston, Canada; Radiology and Nuclear Medicine, CARIM & GROW, Maastricht University, Maastricht, The Netherlands; Department of Radiation Oncology (MAASTRO), Maastricht University, Maastricht, The Netherlands; Department of Molecular Medicine, Aarhus University Hospital, Aarhus, Denmark; Department of Clinical Medicine, Aarhus University, Aarhus, Denmark; Bioinformatics Research Center, Aarhus University, Aarhus, Denmark; Department of Biostatistics, Harvard T.H. Chan School of Public Health, Boston; Princess Margaret Cancer Centre, University Health Network, Toronto, Ontario, Canada; Department of Medical Biophysics, University of Toronto, Toronto, Ontario, Canada; Cancer Research UK Lung Cancer Centre of Excellence, University College London Cancer Institute, London, UK; Cancer Evolution and Genome Instability Laboratory, The Francis Crick Institute, London, UK; Department of Radiology, Brigham and Women’s Hospital, Harvard Medical School, Boston, United States of America

**Author notes:** Joint first authors (Osbert Zalay, Dennis Bontempi). joint last authors (Raymond H Mak, Hugo JWL Aerts). Corresponding author | Hugo Aerts, PhD. Director, Artificial Intelligence in Medicine (AIM) Program, Mass General Brigham & Harvard University Harvard Institutes of Medicine – HIM 343, 77 Avenue Louis Pasteur, Boston, MA 02115. P –, F –.

## Abstract

Because humans age at different rates, a person’s physical appearance may yield insights into their biological age and physiological health more reliably than their chronological age. In medicine, however, appearance is incorporated into medical judgments in a subjective and non-standardized fashion. In this study, we developed and validated FaceAge, a deep learning system to estimate biological age from easily obtainable and low-cost face photographs. FaceAge was trained on data from 58,851 healthy individuals, and clinical utility was evaluated on data from 6,196 patients with cancer diagnoses from two institutions in the United States and The Netherlands. To assess the prognostic relevance of FaceAge estimation, we performed Kaplan Meier survival analysis. To test a relevant clinical application of FaceAge, we assessed the performance of FaceAge in end-of-life patients with metastatic cancer who received palliative treatment by incorporating FaceAge into clinical prediction models. We found that, on average, cancer patients look older than their chronological age, and looking older is correlated with worse overall survival. FaceAge demonstrated significant independent prognostic performance in a range of cancer types and stages. We found that FaceAge can improve physicians’ survival predictions in incurable patients receiving palliative treatments, highlighting the clinical utility of the algorithm to support end-of-life decision-making. FaceAge was also significantly associated with molecular mechanisms of senescence through gene analysis, while age was not. These findings may extend to diseases beyond cancer, motivating using deep learning algorithms to translate a patient’s visual appearance into objective, quantitative, and clinically useful measures.

## INTRODUCTION

Emerging evidence suggests that people age at different rates. Interpersonal differences in genetic and lifestyle factors such as diet, stress, smoking, and alcohol usage have been shown to influence the aging process, and affect DNA methylation status ^1–3^, telomere length ^4–6^, immune and metabolic markers of chronic inflammation, and gene and protein expression patterns ^7–9^. There is no one single clock that measures biological age directly, but establishing biomarkers that correlate with survival time (i.e. time until death) could have clinically relevant applications. Finding an appropriate surrogate of a person’s biological age may provide a better predictor of their physiological health and life expectancy than chronological age. This is especially important in medicine, where both diseases and treatments can cause cellular damage and accelerate the aging process, and an accurate estimation of the biological age could support treatment decisions and allow better quantification of the relative risk-benefit ratio of proposed treatments. For example, a fit 75-year-old whose biological age is ten years younger than their chronological age may tolerate and respond to treatment better and live longer than a 60-year-old whose biological age is ten years older than their physical age.

In current clinical practice, a physicians’ overall impression of a patient constitutes an integral part of the physical exam. It plays a significant role in clinical decision-making in terms of estimating prognosis and weighing the benefits and risks of diagnostic procedures and treatment. However, this is a particularly subjective assessment of functional status or frailty and only a rough estimation of the biological age of a patient ^10^. Especially in oncology, where the therapeutic window is often narrow, and the treatment itself can worsen mortality rates, a decision to treat requires accurately estimating whether the patient would be healthy enough to tolerate treatment and live long enough to benefit from it. Also, we might expect a higher biological age for cancer patients because of the combined impact of the disease and treatment toxicity. Unfortunately, oncologists have to make these complex treatment decisions without knowing the exact biological age of a patient, relying instead on subjective performance status estimates, contributing to a well-documented poor ability to predict the outcome of their patients ^11–13^. Therefore, there is a compelling need for quantitative methods to improve patient stratification and support physicians in this complex decision-making process for appropriate treatment selection. Such an objective measure could also allow for more accurate objective stratification within trials and better translation of results to patients in the real world. Furthermore, this approach could help decipher the biological processes between premature aging and identify individuals that age faster and are at increased risk of diseases.

We hypothesize that a person’s biological age is reflected in their facial characteristics and that deep learning algorithms can capture this information from easily obtainable photographs automatically. Such an approach may provide a more precise measure of a patient’s physiological status than chronological age, providing key information for precision medicine as an actionable clinical biomarker and prognostication factor. Early evidence for this was presented recently by Xia et al. ^14^, in which faces of healthy individuals were measured using a specialized 3-dimensional imaging device, and demonstrated that these data are associated with molecular markers of aging. The authors did not, however, investigate this method in the clinical setting nor its associations with clinical outcomes. Outside of medicine, many academic and commercial institutions are exploring deep learning algorithms to estimate a person’s age and gender from face photographs. The end goal of these efforts is frequently in the realm of marketing and social media to predict end-user habits or preferences ^15–17^, but many datasets and algorithms are readily available and potentially usable for clinical applications.

In this study, we have leveraged recent advances in deep learning algorithms applied to face photographs to estimate a person’s biological age and evaluated its performance in independent clinical datasets from two trans-Atlantic institutions involving a broad spectrum of cancer patient populations (see **Figure 1**). We have shown for the first time that deep learning can estimate biological age from commonly used and easily obtainable face photographs in a clinical context and that this information is associated with medical illness and survival prognostication. Our results demonstrate that face photographs can be used as a novel biomarker source for helping to estimate life expectancy in cancer patients across the disease types and prognoses commonly encountered in an oncology clinic. Furthermore, in end-user experiments, we demonstrate that deep learning-quantified biological age can improve clinician prognostication at the end of life. Our findings motivate the further development of easily translatable deep-learning applications to quantify biological age and inform treatment decision-making for cancer and other medical conditions.

**Fig. 1.**
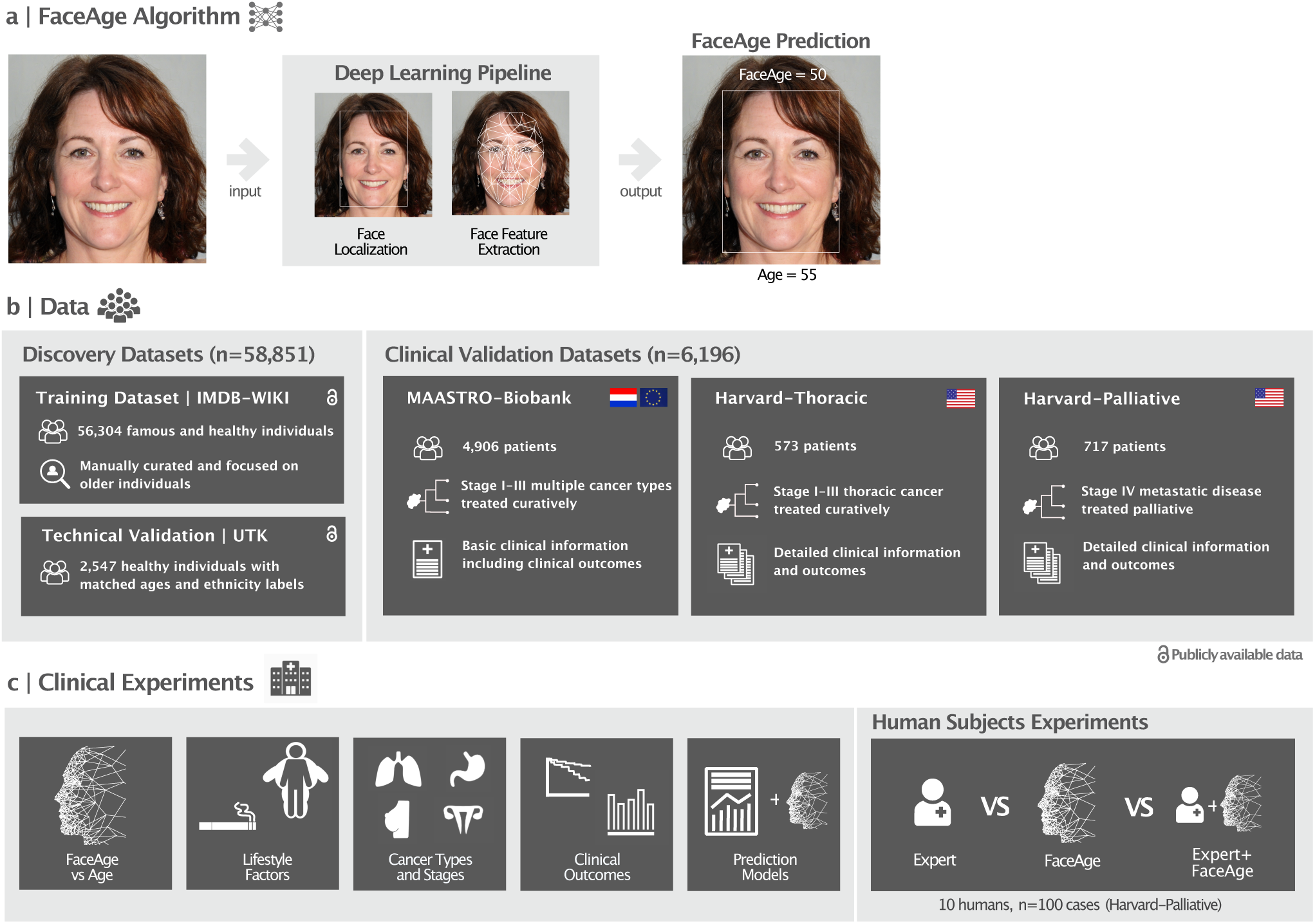
Overview of the study methodology. a) Illustration of the FaceAge algorithm that uses a single photograph of the face as input. First, a convolutional neural network localizes the face within the photograph, and next, a second convolutional neural network quantifies face features and uses these to predict the FaceAge of the person. b) Overview of the datasets used in this study. The FaceAge algorithm was developed using a training dataset with presumed healthy individuals with the assumption that their age closely approximates biological FaceAge. This dataset was manually curated for individuals of 60 years and older to enhance the dataset quality for the age range of the clinical oncology population. The performance of the algorithm was validated across genders and ethnicities in the UTK dataset. Three independent cohorts of cancer patients covering a large spectrum of cancer patients were used to assess the clinical relevance of the algorithm. c) Overview of the clinical experiments performed in this study to assess the clinical utility of FaceAge. The website https://thispersondoesnotexist.com was used to generate the example face photo.

## RESULTS

### FaceAge training and testing across gender and race

We have leveraged recent advances in deep learning algorithms to develop a deep learning system, termed “FaceAge,” to estimate the biological age from a single frontal face photograph as input (see **Fig. 1a)**. First, a deep learning network automatically localizes the face within the picture; next, a second deep learning network uses this extracted image as input to estimate the age. The deep learning system was trained and tested to predict age in a dataset of 56,304 presumed healthy individuals (particularly politicians, actors, and other well-known people). We assume that the people included in this cohort are of average health (i.e., having chronological age close to their biological age). This dataset was manually curated and an older chronological age range was selected for quality assurance, reflecting the clinical oncology population (see **Fig. 1b**). As a technical validation, we evaluated the performance of FaceAge on face photographs obtained from an independent dataset, UTKFace (UTK) (*n* = 2,547) ^42^. We found comparable performance in all gender and ethnic groups with chronological age being close to FaceAge (see appendix p. 5).

### Illness and Lifestyle factors influence FaceAge

To assess the clinical relevance of FaceAge estimates in patients with a cancer diagnosis, we performed detailed experiments in three separate clinical cohorts from two institutions (see **Fig. 1b-c**; appendix p. 10-11). All patients had a face photograph acquired before radiation treatment as part of the routine clinical workflow. We found that cancer patients had a significantly higher FaceAge than chronological age (*n* = 6,367, mean increase of 4.79 years, paired two-sided t-test, *P* < 0.001, **Fig. 2a**), indicating that cancer patients, on average, look older compared to their age. This was consistent across cancer types. This contrasted with the results in the presumed healthy populations. Firstly, in the UTK validation dataset, we found a significantly smaller difference between FaceAge and age (mean increase of 0.35 years) compared to cancer cohorts (unpaired two-sided t-test, *P* < 0.001), indicating that individuals in the general population look more similar to their age, as expected. Additionally we analyzed the faces of patients treated for benign conditions, as well as ductal carcinoma in-situ (DCIS) patients, with DCIS being a precancerous condition of the breast in which approximately 30% of patients go on to develop invasive breast cancer if left untreated. The non-cancerous cohorts had a smaller FaceAge-to-age gap in comparison to cancer patients (median difference 3.41 years compared to 4.55 years for cancer patients, *P* < 0.001), with the benign patients having FaceAge closest to chronologic age (median difference 1.95 y compared to cancer patients, *P* < 0.0001), and DCIS patients having intermediate FaceAge values (median 3.86 y compared to cancer patients, *P* = 0.019) (see appendix p. 6), further supporting the claim that patients with cancer look older compared to individuals without cancer.

**Fig. 2.**
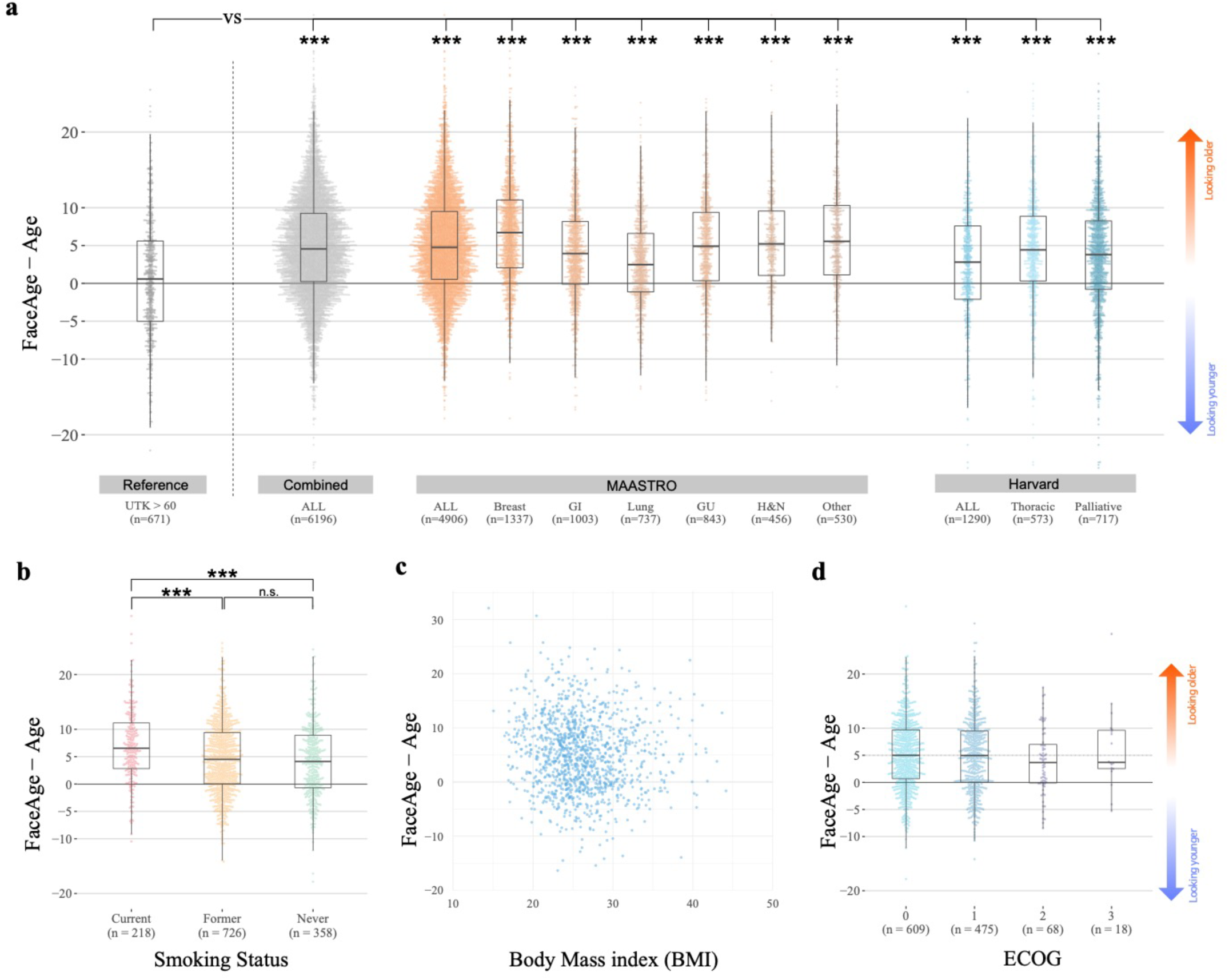
Application of FaceAge in cancer patients. a) Difference between FaceAge and age across cancer types and datasets to investigate if individuals look older or younger compared to their age. Analyzing all cancer patients included in our analysis, we found that, on average, cancer patients look older than their age. This difference was significant when comparing these results to the reference UTK dataset of healthy individuals 60 years and older. b) Difference between FaceAge and age for current, former, and never smokers included in the MAASTRO cohort. c) Scatterplot assessing the association of FaceAge with body mass index (BMI) in the MAASTRO cohort. d) Association of FaceAge with performance status (Eastern Cooperative Oncology Group (ECOG)) was quantified for a subset of patients in the MAASTRO cohort. In the boxplots the box covers the interquartile range (IQR), with the line at the center of the box indicating the mean; the top (bottom) whisker extends from the box to the largest (smallest) value within 1.5 IQR (***: p<0.001; unpaired, two-sided t-test was used in all cases).

To assess the effect of lifestyle factors, we compared the difference between FaceAge and age in current, former, and never smokers in the MAASTRO cohort. We found that current smokers look significantly older (mean increase = 33.24 months; unpaired two-sided t-test, t = 4.78, 95% CI 1.63-3.91, *P* < 0.001) compared to former and never smokers (see **Fig. 2b**), which was consistent across cancer types (appendix p. 7). Interestingly, we did not find a significant difference between former smokers and never smokers (mean increase = 5 months; unpaired two-sided t-test, t = 0.96, 95% CI −0.44-1.28, *P* = 0.34), indicating there may be a reversible effect of smoking on facial aging characteristics. To assess the effect of weight, we compared FaceAge with body mass index (BMI) (see **Fig. 2c** and appendix p. 7). Although a statistically significant association (*n* = 1295; *r* = −0.0999; *P* < 0.001) was observed, the effect size was minimal, indicating only a weak relationship between FaceAge and body mass index. As the Eastern Cooperative Oncology Group (ECOG) performance status is used for clinical stratification, we compared the association of ECOG groups with the difference between FaceAge and age (see **Fig. 2d** and appendix p. 7). In both the MAASTRO and Harvard cohorts, we found no statistically significant differences (unpaired two-sided t-test, *P* > 0.092) between the groups, indicating that FaceAge quantifies different biological information relative to the performance status of a patient.

### FaceAge is a biomarker for longevity across a wide spectrum of clinical settings

To assess the prognostic relevance of FaceAge estimation, we first assessed how FaceAge predictions associated with survival. The MAASTRO cohort contains a broad population of patients with a variety of non-metastatic cancer types and a wide range of prognoses (*n* = 4,906, median age = 67 [range = 22-94] years; median survival = 36.0 months). Kaplan-Meier survival analysis revealed good stratification of increasing mortality risk with increasing FaceAge risk groups (see **Fig. 3a**). In univariate analysis, all FaceAge risk groups showed significantly worse survival than the youngest-looking FaceAge risk group. This result remains significant after adjustment for age, gender, and tumor site for the two oldest-looking risk groups (see **Fig. 3b**). This was confirmed by assessing FaceAge as a continuous parameter, which demonstrated significant prognostic performance (*P* = 0.0013) after adjusting for age, gender, and tumor site in the whole cohort (see **Fig. 3c** and appendix p. 12). Analyzing specific cancer types, we found that FaceAge was significantly predictive in all cancer sites, which remained significant after correcting for age and gender for patients with breast cancer, genitourinary cancer, and gastrointestinal cancer (*P* < 0.05; see **Fig. 3c**).

**Fig. 3.**
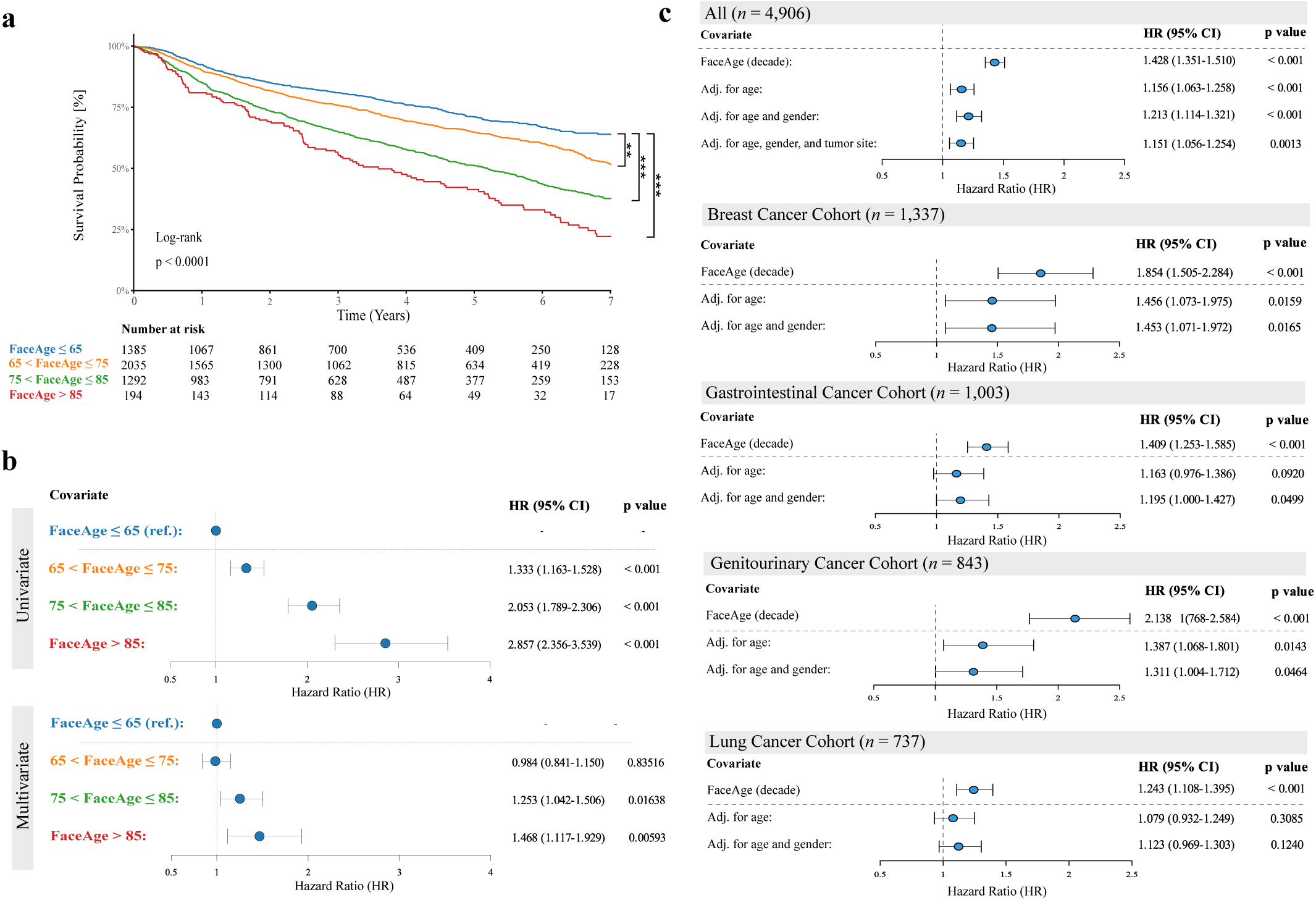
Prognostic performance of FaceAge in Several cancer cohorts. a) Kaplan-Meier survival analysis shows significantly worse survival for increasing FaceAge estimation from only a face photograph as input. b) Forest plots of FaceAge risk groups demonstrate significant differences for all groups univariately, and this remains significant after adjustment for age, gender, and tumor site for the two oldest-looking groups compared to the youngest group as reference. c) Forest plots of FaceAge estimates as a continuous parameter for all patients as well as the four largest tumor sites. Note that FaceAge is significant in all tumor sites in univariate analysis and remains significant in breast, gastrointestinal, and genitourinary cancer after correction for age and gender. HR: Hazard Ratio; CI: confidence interval; (**: P < 0.01; ***: P < 0.001; all analyses are performed in the MAASTRO Cohort, n = 4,906)).

Next, we evaluated FaceAge in the Harvard-Thoracic cohort, a site-specific dataset with thoracic malignancies (*n* = 573, median age = 69.0 [33.3-93.2] years; median survival = 16.9 months), of which the majority were non-small cell lung cancer (NSCLC) patients (n=450, 78.5%). Granular clinical data were available for these patients, which allowed for further investigation into the independent performance of FaceAge. Therefore, we investigated key clinical factors known to affect survival in lung cancer, including clinical stage, ECOG performance status, smoking history, gender, histology, and treatment intent. Although in univariate analysis, FaceAge was not significant, in multivariate analysis, FaceAge became statistically significant after adjusting for these clinical factors (per decade HR 1.15, 95% CI 1.03-1.28, *P* = 0.0113, appendix p. 13). Adding FaceAge to the base model also added explanatory value (By doing incremental sensitivity analysis through the addition of individual covariates to the univariate model, the significance of FaceAge was unmasked by adjusting for Stage I lung cancer patients in particular (incrementally adjusted per decade HR 1.17, 95% CI 1.05-1.29, *P* < 0.003). FaceAge did not appear to be prognostic for these very early-stage patients, although other competing risk factors for death, such as comorbidities, could have a greater impact on this subgroup and account for the lack of prognostic power (e.g. many early-stage lung cancer patients receiving radiotherapy are not surgical candidates due to comorbidities, which is the gold standard treatment for early-stage disease). Comparing these results to age, we found that chronological age was neither significant on univariate analysis nor after adjustment for multivariable clinical factors (per decade HR 1.08, 95% CI 0.97-1.21, *P* = 0.162). Furthermore, we observed a significant increase in model explanatory power when adding FaceAge to the multivariate model (log-likelihood ratio test (LLR), chi-squared statistic [1 deg freedom]: 6.501, *P* = 0.0108), while this was not observed when adding chronological age (LLR test, chi-squared statistic [1 deg freedom]: 1.965, *P* = 0.161). These results show that FaceAge consistently improves prognostication while age does not, and that FaceAge contains prognostic information that is not captured by other investigated clinical parameters.

### FaceAge can improve the performance of clinical models

As a direct and relevant clinical application of FaceAge, we assessed the performance of FaceAge in end-of-life patients with metastatic cancer that received palliative treatment. In these patients, clinical prediction models can help improve physicians’ decision-making as to whether or not to administer treatment, as well as the appropriate treatment intensity, both of which are largely a function of a physician’s impression of overall prognosis, performance status, and frailty. We assessed the independent prognostic performance of FaceAge in the Harvard-Palliative dataset (*n* = 717; median age = 65.2 years; median survival= 8.2 months). Covariates that are known to be related to survival in palliative cancer patients ^27,43^, such as performance status, number of hospitalizations and emergency room visits in the past three months, sites of metastatic disease, and the primary cancer type, were analyzed. FaceAge was found to be significant in both uni– and multivariate analysis (univariate per decade HR 1.10, 95% CI 1.01-1.21, *P* = 0.035; multivariate HR 1.12, 95% CI 1.02-1.23, *P* = 0.021), whereas chronological age was not significant in either uni– or multivariate analysis (see appendix p. 14-15). We also observed a significant increase in explanatory power by adding FaceAge to the multivariate model (LLR test, chi-squared statistic [1 deg freedom]: 5.439; *P* = 0.020), which was not observed when adding chronological age (LLR test, chi-squared statistic [1 deg freedom]: 2.548; *P* = 0.11).

Next, we evaluated the additive performance of FaceAge to a clinically validated risk-scoring model for palliative patients. This TEACHH model ^27^ was originally developed to estimate the survival time of palliative cancer patients by using a clinical risk-scoring system based on six covariates: type of cancer, ECOG performance status, presence of liver metastasis, number of prior palliative chemotherapy courses, age at treatment, and number of previous hospitalizations. In the Harvard-Palliative cohort, this model demonstrated significant performance to stratify patients in different risk groups. If we substituted chronological age with FaceAge, we found this significantly increased the model’s discriminatory power, as quantified by a significantly increased log-likelihood ratio when comparing the three risk categories of the TEACHH model against the baseline hazard for FaceAge (LLR test, chi-squared statistic [2 deg freedom]: 75.1; *P* < 10^-16^), compared to age (LLR test, chi-squared statistic [2 deg freedom]: 63.3; *P* < 10^-13^). This was also reflected in better separation of risk groups by survival, as substituting FaceAge lowered the median survival (MS) and increased the hazard ratio (HR) of the highest risk group (FaceAge high-risk group: MS 2.5 months, HR 2.74, *P* < 0.001; chronologic age high-risk group: MS 2.9 months, HR 2.43, *P* < 0.001), and raised median survival and decreased HR of the lowest risk group (FaceAge low-risk group: MS 2.2 years, HR 0.22, *P* < 0.001; chronologic age low-risk group: MS 1.9 years, HR 0.28, *P* < 0.001).

### FaceAge supports physicians’ clinical decision-making ability at the end-of-life

To compare FaceAge with the ability of humans to predict the overall survival of metastatic cancer patients, we performed a survey using 100 cases randomly drawn from the Harvard Palliative cohort. First we assess the performance of humans for estimating six-month survival from only face photographs, without the benefit of additional clinical information, by asking ten medical and research staff members at Harvard-affiliated hospitals (5 attending staff physicians who were all oncologists or palliative care physicians, 3 oncology residents, and 2 lay [non-clinical] researchers), to predict whether the patient would be alive at 6 months (an important endpoint to guide decision-making at the end-of-life). Here, we found that attending physicians performed the best overall, while there was a notable performance difference between individuals within each group (**Fig. 4a**). This was also shown by Kaplan-Meier analysis, demonstrating that the highest-performing attending physician predicting six-month survival was able to stratify patients into high and low-risk groups that demonstrated significant survival differences (median survival: high-risk group 4.8 months, low-risk group 13.2 months, log-rank test, *P* < 0.005), whereas the lowest-performing physician did not (median survival: high-risk group 7.7 months, low-risk group 13.2 months, *P* = 0.49) (**Fig. 4b**).

**Fig. 4.**
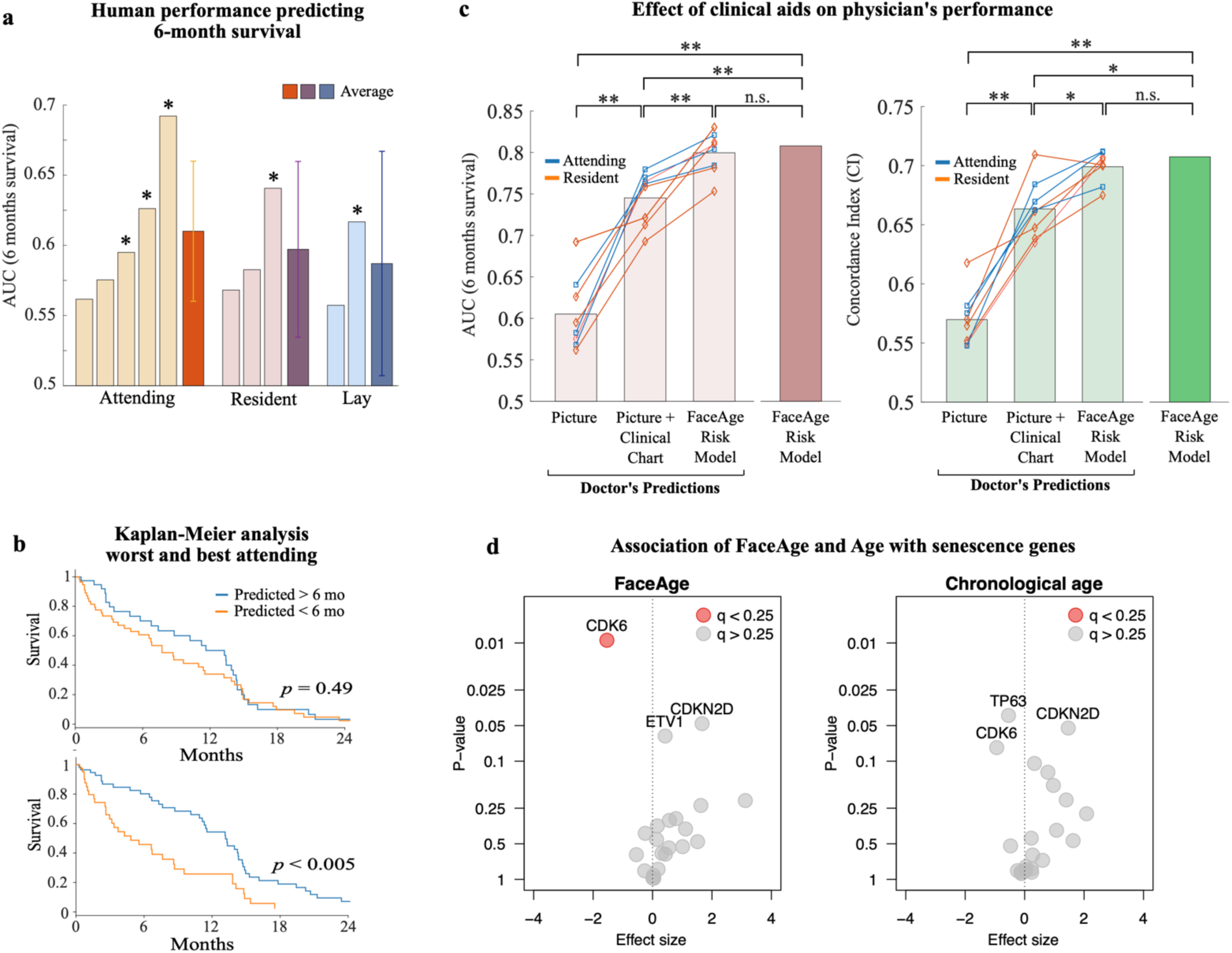
Comparison of human and FaceAge performance predicting survival. a) Area under curve (AUC) of the receiver operating characteristic for 6-month survival predicted for 10 survey takers, grouped by experience level (*: AUC significantly different from random). Confidence intervals are shown for average AUCs. b) Kaplan-Meier analysis of overall survival of patients predicted to be either alive at 6 months or not, comparing lowest (top) and highest (bottom) performers of the attending physicians. c) Six-month survival prediction (left) and overall survival time (right) for physicians (both attending and residents) aided with only a picture, a picture + clinical chart information, and a risk model including clinical data and FaceAge. (two-sided Wilcoxon signed rank test; *: p<0.05; **: p<0.01; n.s.: non-significant). The survey included 100 palliative patients randomly selected from the Harvard-Palliative cohort. d) Results of the Burden test for the association of the senescence genes with FaceAge or chronological age. After adjusting for multiple comparisons using a false discovery rate (q) of 0.25, only CDK6 was statistically significant for FaceAge, while none of the other genes were statistically significant for either FaceAge or chronological age.

Next, to evaluate the complementary value of FaceAge with other clinical data (primary cancer diagnosis, age at treatment, performance status, location of metastases, number of emergency visits, number of hospital admissions, prior palliative chemotherapy courses, prior palliative radiotherapy courses, time to first metastasis, and time to oncology consult), we trained a FaceAge Risk Model, which combined the clinical factors with FaceAge to predict survival probability. In successive survey rounds, we asked the clinical survey-takers to predict six-month survival based on a face photo alone, the face photo provided together with the patient clinical chart information, and then with the addition of the FaceAge Risk Model (**Fig. 4c**). Here, we found that human performance significantly increased (*P* < 0.01) if we provided face photographs combined with clinical chart information (AUC = 0.74 [0.70-0.78]), compared to a face photograph only (AUC = 0.61 [0.57-0.64]). However, human performance was improved even further (*P* < 0.01) when the FaceAge Risk Model was made available to clinicians in addition to chart information (AUC = 0.80 [0.76-0.83]), with the latter performance not being statistically different (*P* = 0.55) from the FaceAge Risk Model alone (AUC = 0.81 [0.71-0.91]). Similar results were found for overall survival as quantified by the concordance index.

In general, consensus between doctors and risk models was high, and performance accuracy was good for the straightforward cases, whereas consensus and accuracy suffered in the more challenging cases with low observer agreement. The best individual performance was seen with the inclusion of the FaceAge risk model, which helped convert some of the clinicians’ inaccurate predictions into accurate ones.

### FaceAge demonstrates association with senescence genes

To evaluate whether FaceAge has the potential of being a biomarker for molecular aging, we performed a gene-based analysis to measure its association with senescence genes in comparison with chronological age. The analysis was conducted on 146 individuals from the Harvard Thoracic Cohort who were diagnosed with non-small cell lung cancer and profiled using whole-exome sequencing. We evaluated 22 genes known to be associated with senescence (appendix p. 9), and we found that FaceAge was significantly associated with CDK6 after adjusting for multiple comparisons (false discovery rate of 0.25) (**Fig. 4d**). CDK6 has an important role in regulating the G1/S checkpoint of the cell cycle through phosphorylation and activation of the Rb (retinoblastoma) tumor suppressor protein by complexing with CDK4 and Cyclin D. By contrast, no genes showed a significant association with chronological age after adjustment for multiple comparisons. While limited in scope to only a small set of preselected genes to conserve statistical power, this analysis illustrates the potential of using FaceAge to discover associations with genes related to biological aging, which are different from and may not be detected by chronological age.

## DISCUSSION

The results of our work demonstrate that facial features captured in a photograph contain prognostic information related to the apparent age of a person, informing survival predictions in cancer patients. Our methodology, which relies on easily obtainable face photographs, improves upon the current standard of subjective visual assessment by clinicians. Moreover, we show for the first time that a deep learning model trained for age estimation on a healthy population can be used clinically to stratify sick patients according to survival risk based on their facial appearance, and that this novel biomarker is more predictive than patient chronological age. We found that cancer patients, on average, look approximately five years older than their stated age, and also have statistically higher FaceAge compared to clinical cohorts of non-cancer patients treated for conditions that are benign or precancerous. FaceAge was prognostic of survival time and provided additional explanatory power compared to chronologic age, even when FaceAge was adjusted for chronologic age in the same survival model. However, adjustment for chronologic age expectedly reduces the effect size (i.e., hazard ratio) of FaceAge, given the co-dependence, so the most appropriate way to deploy FaceAge in risk models is to substitute it for chronologic age. FaceAge outperformed age in univariate and multivariate analyses across several cancer sites and clinical subgroups, even after adjusting for known clinical risk factors. Notably, FaceAge performed well both in patients treated for curative intent, with life expectancies of several years, as well as in patients at end-of-life with an expected survival of weeks to months. We also demonstrated that FaceAge significantly improved the performance of a validated clinical risk-scoring model ^27^ for estimating survival in end-of-life patients who received palliative radiation treatment, a patient population for which there is a critical need to improve treatment decision-making utilizing such models. We showed clinician survival prediction performance improved when FaceAge risk model predictions were made available to them, especially among physicians with lower baseline performance. Lastly, we provided evidence from SNP gene analysis that FaceAge is correlated with molecular processes of cell-cycle regulation and cellular senescence, supporting the hypothesis that FaceAge is a biomarker that relates to biological aging, consistent with its interpretation as a modifier of survival time in a diseased population. We also found an inverse association of FaceAge with skeletal muscle, a known prognostic factor for mortality in cancer patients ^44,45^, indicating a link between FaceAge and patient frailty.

We selected oncology as the clinical focus of this paper because these patients are closely followed in terms of their survival outcomes, and their disease processes and treatments can significantly impact biological aging, with traditional clinical decision-making relying on clinicians’ largely subjective estimates of whether the patient would be fit enough to tolerate treatment. We further demonstrated the clinical applicability of our methodology by evaluating FaceAge over a wide spectrum of cancer types and stages commonly encountered in an oncology clinic, but with a focus on oncology patients who received radiotherapy, because, unlike other patients, these patients routinely have their face photograph taken as part of the treatment registration process.

As clinicians currently have to base their decisions on scant information about the true biological age of a patient, relying instead on subjective measures such as performance status (e.g., Eastern Cooperative Oncology Group or Karnofsky performance score, usually ranked by a health care provider after obtaining verbal history about the patient’s level of functioning or coping, a task fraught with inter-rater reliability issues, reporting and recall bias) there is a compelling need for better biomarkers to estimate biological age. In turn, improved estimation of biological age can inform survival prediction, a very challenging task even among experienced oncologists ^43^. Indeed, clinicians are often not comfortable predicting and disclosing life expectancy ^46^, and are systematically over-optimistic in their prognoses ^11,46–48^. We showed that FaceAge predictions improve oncologists’ ability to accurately estimate survival, which can help support patient-centered care by improving the accuracy of information provided during patient counseling, but even more crucially, it can help guide treatment decisions. This is not to say that the predictions of the FaceAge model should be used in isolation to make important life-and-death medical decisions, which would be ill advised. Rather, the purpose is to improve predictive power and accuracy of existing clinical risk models, whose power lies in combining aggregate clinical covariates so that the resulting outcome predictions are considerably more robust. To that end, we have studied and demonstrated the highly clinically impactful application of FaceAge in predicting survival at end-of-life in metastatic cancer patients by combining the model predictions with the covariates of the already-validated TEACHH model. Unnecessarily aggressive end-of-life medical interventions can negatively impact quality of life and increase health care expenditures, and prognostication with FaceAge may help clinicians provide their patients with more accurate prognostication to make better-informed decisions about care for a variety of end-stage diseases.

Practical benefits of our methodology include easy implementation in the clinic. Our model can draw upon data from pre-existing electronic medical records available to the clinician, and a biological age estimate and related outcomes prediction can be computed in near real-time using a standard desktop computer or smartphone. Face photos are readily obtainable under most circumstances, and multiple images can be acquired before, during, and after treatment, and can be stored without using significant digital resources. Protocols can be implemented to standardize how digital face images are obtained so that the process can be rendered robustly reproducible. Even phone cameras may be utilized for image capture making accessibility straightforward. Some major electronic health record systems already have the capacity to directly and securely upload and store photographs taken on digital cameras, including phone cameras.

Early evidence supporting our main hypothesis that biological aging is reflected in facial characteristics was presented in the recent study by Xia et al. ^14^ In this study, the authors used a specialized 3-dimensional imaging device to obtain morphological face images of healthy individuals, and demonstrated that deep learning networks could infer molecular markers of aging based on lifestyle and dietary factors. They did not, however, investigate any direct association with clinical outcomes, and the requirement for specialized equipment used in their study might limit the widespread application of such a methodology. Our approach, which uses standard face photographs, provides an important improvement with respect to applicability of such methods in the clinical setting and beyond. Although many academic and commercial applications for estimating age and gender from face photographs have been explored, the end goal of these efforts frequently are tied to marketing and social media in order predict end user habits or preferences, whereas our objectives are clinically focused. We are the first to demonstrate the utility of face photographs as a novel biomarker source in medical applications by its association with medical illness and survival prognostication.

A major strength of our study is that the FaceAge model was entirely trained on publicly-available, non-clinical databases. Such databases have the advantage of their large size and accessibility over smaller, more limited institutional datasets, thereby making the model more usable in a real-world setting. Institutional datasets tend to be highly restricted in terms of access, despite the benefit of face photographs being associated with clinical health parameters. The standard approach to training deep-learning systems is to train a model on datasets that are very similar to the operational datasets. Although this may improve performance on the operational dataset, it comes at the expense of limited generalizability. Such models tend to be brittle in that they fail when applied on new datasets with differing characteristics from the training set. However, the performance of the FaceAge model explicitly relies on a presumed difference between healthy and diseased populations, with the hypothesis that the predicted age differential reflects a component unrelated to model error but is instead attributable to the intrinsic difference between age and biological age. The fact that stratification of this age difference has prognostic value in terms of identifying higher– and lower-mortality risk groups of cancer patients in our study, drawn from three separate, heterogeneous datasets originating from two trans-Atlantic institutions, supports not only our hypothesis, but also the generalizability of our method, which is remarkably agnostic to disease site, treatment type, and disease severity with respect to cancer patients.

The training data we used do contain expected limitations and possible intrinsic biases. Firstly, as mentioned above, the online image databases used for training lack associated health information. We made the implicit assumption that the subjects in the training data were of average health for their age (that is, they have a biological age similar to their age), though this is clearly not true in all cases. Moreover, the images contain a significant proportion of public individuals and well-known figures such as film actors and politicians, which in and of itself might introduce a systematic biological age selection bias, because such individuals might have a different biological age compared to an age-matched cohort of “non-famous” peers, due to different lifestyle and socioeconomic factors. Actors in particular might have other cosmetic or facial alterations that could affect biological age estimation from such photographs, in addition to more frequent digital image touch-ups. While we do not have statistics on how many of the photos included cosmetic alterations of the physical or digital variety, it is unlikely that it would represent a significant proportion of the whole, as the IMDb-Wiki database contains many photos of people other than actors, including writers, philanthropists, educators, scientists and people from all domains of society. As well, the large size of the heterogeneous dataset tends to average out potential biasing factors. Because the deep learning network is trained to predict the chronological age of a large cross-section of individuals with no expected consistent age-dependent pattern of facial alterations, the network would be anticipated to ignore such variations when forming pattern associations with age. Furthermore, photos in the wild (i.e. the public domain) are non-standardized and heterogeneous, meaning differences in image sizes, resolution, numbers of people in the image, backgrounds, clothing worn, and other photographic artifacts or distortions will be present in many of the training images. Due to the large size of the training dataset, combined with our manual image quality assurance, the network training was able to successfully account for these artifacts. Indeed, we demonstrated the robustness of the model to image perturbations using sensitivity analysis. Moreover, the noisiness of the training data even likely improved the generalizability of our model, as demonstrated by the FaceAge model’s significant performance in the real-world clinical datasets.

The use of facial photographs in our analyses presents multiple ethical considerations. Perhaps the most obvious is that facial photographs are uniquely identifying, and in the case of their use in medical charts, tied to sensitive personal health information. Although a model trained both on patient face photographs and patient chart data might demonstrate better predictive performance than one trained on publicly available face images, sharing both the training data and the derived models could lead to the ability to re-create identifying face images through backwards-inferring the original training datasets. For such a model, clinical deployment for healthcare-related outcomes prediction would necessitate a secure institutional setting using appropriate privacy protocols that are similar to those already in place for other sensitive electronic medical records. Another concern is that patients may not realize their face photograph is part of the medical record, and therefore may not have an expectation that these images would be used to inform survival prediction, raising potential concerns with regard to informed consent and secondary use of data. In addition to infringement of patients’ autonomy and right to privacy, inappropriate implementation of such a model could have unforeseen consequences including impacting the physician-patient relationship and affecting care decisions beyond the use cases studied here. Therefore, it is imperative to clearly communicate to patients the specific clinical goals and reasons for using such face image data, and obtain explicit informed consent before these data are used in the clinical setting. Finally, the intended usage of the model should be published alongside the model itself, and the model’s application in clinical or research settings should not extend beyond the defined scope.

Outside of healthcare, it is possible to envision several potential misuses of such a model. These include health, disability, and life insurance payors incorporating estimated survival metrics from face images to determine the insurability of prospective policy holders, or a technology or media company promoting health or lifestyle products with targeted advertising based on client biological age estimation. Strong regulatory oversight would be a first measure toward mitigating this problem. Another important ethical concern is racial or ethnic bias, which has been problematic for automated face recognition software, especially in legal and law enforcement applications ^49^. The potential for racial bias is addressed in our model in several ways: Firstly, we demonstrated that the model age predictions are fairly balanced across different ethnic groups drawn from the UTK validation dataset, which is one of the most ethnically diverse age-labeled face image databases available publicly and therefore appropriate for assessing model performance in this regard, with non-Caucasian individuals comprising approximately 55% of the database ^50^; secondly, ethnicity was treated as a covariate that we adjusted for in multivariable analysis of the clinical datasets, which revealed that the FaceAge measure was minimally impacted by ethnicity. Our model is configured for the task of age estimation, which in our opinion has less embedded societal bias than the task of face recognition ^51^. However, it will be important to continue to assess for bias in performance across different populations, as differential treatment decision-making as a result of its predictions could amplify existing health disparities. Institutional and governmental oversight of how such models are regulated and deployed, with careful prescription of their intended use, and educational support for clinical end-users (including appropriate use case and model failure modes) will be crucial to ensure that patients can benefit from their incorporation into clinical care while minimizing the risk of abuse, unintended or otherwise.

Future work involving FaceAge will focus on improving the predictive power and accuracy of the methodology, broadening it to other applications within and beyond oncology. As face photographs can easily be obtained, serial data collection prior, during, and after treatment should be incorporated into the model to assess how FaceAge varies over time, and what factors might be driving these changes. Incorporating other photographic or videographic image data beyond the face to enable characterization of body morphology, posture, skin texture, presence of edema, etc. might lead to more accurate biological age estimation and improve model predictive power.

Furthermore, these measures could be combined with other known and emerging biological correlates of aging, including molecular markers (obtained from the peripheral blood, for example ^52,53^), gene expression ^8^, body composition ^54^, microbiome ^55^, and other imaging modalities such as x-rays ^56^, CT ^57^ or MRI ^58^. We have already demonstrated that FaceAge is correlated with genes implicated in cellular senescence, as well as with skeletal muscle index, a body composition measure already known to be prognostic of survival in cancer patients ^44,45^. By combining independent predictive markers together with FaceAge, it may be possible to provide a more holistic and accurate view of a person’s aging process, because different tissues, organ systems and individuals age at different rates in a manner that is complex and nonlinear, dependent on multiple interacting factors that are both internal and external.

Extending the method to other patient populations outside of oncology (for example, assessing whether a diabetes patient would benefit from a new anti-hyperglycemic agent, or whether an elderly orthopedic patient would tolerate hip replacement surgery) should be explored, because biological age would be expected to play a role in modulating the impact of a wide range of disease processes. Estimation of biological age could provide a time-dependent window on health status, which can fluctuate and even reverse course, while chronological age is a monotonically increasing function of time regardless of health status. Therefore, it may be possible to use FaceAge or similar biological age markers to track the effect of lifestyle interventions. By stopping smoking, starting to exercise, or changing diet, for example, it may be possible to quantify the slowing or reversal of biological aging.

In conclusion, we have demonstrated that a deep learning model can enhance survival prediction in cancer patients through analysis of face photographs via estimating patients’ biological age from their facial features. We have demonstrated that deep learning-based FaceAge estimates are prognostic in a wide range of cancer types and clinical settings, and can be integrated with existing clinical chart information and clinical risk-scoring models to improve clinicians’ prediction performance. Further research and development must be carried out, however, before this technology can be effectively deployed in a real-world clinical setting.

## MATERIALS AND METHODS

### Datasets

A detailed description of the datasets used in this study can be found in appendix p. 2-3.

### FaceAge Deep Learning Pipeline

The FaceAge deep learning pipeline comprises two stages, a face location stage and a feature encoding stage. The first stage pre-processes the input data by locating the face within the photograph and defines a bounding box around it, then crops the image around the face, resizing and normalizing it. The second stage takes the extracted face image and feeds it into a convolutional neural network (CNN) that encodes image features, which through regression yield a continuous FaceAge prediction as the output. The face localization stage utilizes a fully-trained multi-task cascaded convolutional neural network developed by Zhang et al ^18^. The network is composed of three sub-networks, namely a proposal network (P-net) that creates an initial set of bounding box candidates, of which similar boxes are merged then further refined (R-net) using bounding box regression and face landmark localization, then the third stage (O-net) makes more stringent use of face landmarks to optimize the final bounding box, achieving a test accuracy of 95%. Once the face has been extracted from the photograph, the image is resized to a standard set of dimensions and pixel values normalized across all RGB channels. The feature encoding stage makes use of the Inception-ResNet v1 architecture ^19^, pre-trained on the problem of face recognition ^20,21^. The convolutional neural network creates a 128-dimensional embedding vector as a low-dimensional representation of face features. To develop a model for age regression, a fully-connected output layer was included that uses a linear activation function. Transfer learning was then applied to tune the weights of the last 281 of the 426 Inception-ResNet layers (Inception Block B1 onward) in addition to the fully-connected output layers, using the augmented and randomly rebalanced training dataset of age-labeled face images derived from the IMDb-Wiki database. Training was carried out on paired GPUs using Keras with Tensorflow backend, applying stochastic gradient descent with momentum for backpropagation, minimizing mean absolute error (MAE), with batch size of 256, batch normalization, dropout for regularization, and initial learning rate of 0.001 with incremental reduction on plateauing of error rate. The model development set was subdivided using random partitioning into 90% training, 10% testing. Model performance was good for the clinically relevant age range (60 years or older) that underwent manual curation and quality assurance (MAE = 4.09 years).

### Statistical and Survival Analysis

Independent model validation was carried out by examining age estimation performance on the publicly-available UTKFace dataset (see appendix p. 5), and on clinical datasets from MAASTRO and Harvard by comparing FaceAge predictions for non-cancerous clinical cohorts to predictions from the oncology clinical datasets (see appendix p. 6). Statistical and survival analyses were performed in *Python* 3.6.5 using the Lifelines library, as well as NumPy and SciPy libraries, and in the open-source statistical software platform, *R*. The clinical endpoint was overall survival (OS). Actuarial survival curves for stratification of risk groups by overall survival were plotted using the Kaplan Meier (KM) approach ^22^, right-censoring patients who did not have the event or were lost to follow-up. All hypothesis testing performed in the study was two-sided, and paired tests were implemented when evaluating model predictions against the performance of a comparator for the same data samples. Differences in KM survival curves between risk groups were assessed using the logrank test ^23^. Univariable and multivariable analysis via the Cox proportional hazards (PH) model ^24^ was carried out to estimate and adjust for the effect of clinical covariates such as gender, disease site, cancer stage, smoking status, performance status, treatment intent, time to treatment as well as the number of courses of radiotherapy and chemotherapy in non-curatively treated patients. Effect size is related to the magnitude of the hazard ratio (HR), and confidence intervals for the hazard ratios in Cox univariate and multivariate regressions were computed to estimate the uncertainty in the effect size of the covariates. Compared to the Harvard Thoracic and Palliative datasets, the MAASTRO dataset has far fewer covariates with which to adjust the model, limited to age, gender, and cancer site (primary diagnosis). However, due to the large sample size of the MAASTRO dataset compared to the smaller Harvard datasets, there was sufficient statistical power to investigate FaceAge and age variables together in the same model for multivariate analysis in order to determine if FaceAge had additional prognostic information that was complementary to age. The correlation coefficient between FaceAge and age was below 0.75, so we did not include an interaction term in the Cox PH analysis (see appendix p. 12). To assess the change in the explanatory power of a model by the addition of a study variable (e.g., FaceAge, age, etc.), as well as overall model fit, the log-likelihood ratio test was utilized ^24^. The robustness of survival predictions was assessed by the concordance index ^24,25^ and by the area under the curve (AUC) of the receiver-operating characteristic (ROC) ^25,26^. For the Harvard Thoracic cohort, BMI was excluded from the multivariate analysis due to the small number (*n =* 106) reporting on that covariate. For the Harvard Palliative, a forward selection threshold of 0.2 for the *P*-value was used, to avoid overfitting the model due to the large number of possible covariates in relation to the number of events.

In addition to univariate and multivariate analysis, for the Harvard Palliative treatment cohort, the performance of FaceAge was compared directly to age by substituting one variable for the other in a clinically validated risk-scoring prognostic tool for estimating life-expectancy of palliative cancer patients treated with radiotherapy, called the TEACHH model ^27^. The risk-scoring tool was developed by Krishnan et al. to identify which patients might benefit from palliative radiotherapy (i.e. those predicted to survive longer than 3 months). All TEACHH model variables except age were kept the same (type of cancer, Eastern Cooperative Oncology Group (ECOG) performance status, prior palliative chemotherapy, prior hospitalizations, and presence of hepatic metastases) in order to enable fair comparison of the effect of substituting FaceAge in place of the age. Scoring rules were kept identical to the original TEACHH model. By adding up the score, the individual is stratified into one of three risk categories (A: low risk (1-2 points), B: medium risk (2-4 points), C: high risk (5-6 points)), with highest and lowest risk groups predicting for survival of 3 months or less, versus 1 year or greater, respectively, which are clinically relevant survival endpoints for palliative patients. We performed Kaplan-Meier analysis on the respective risk models incorporating FaceAge versus age, overlaid to show the relative change in actuarial survival curves, and quantified the differential contribution of FaceAge to the explanatory power of the TEACHH model compared to age, using the log-likelihood ratio, and by computing the relative change in magnitude of the hazard ratios for the TEACHH model’s three risk categories (see appendix p. 8).

### Establishing evidence for FaceAge as a biomarker for molecular aging

We evaluated the association of single-nucleotide polymorphisms (SNPs) with FaceAge or chronological age by running a gene-based analysis. Lymphocyte DNA from blood samples were collected and whole exome sequencing was conducted using the Illumina Infinium CoreExome Bead Chip. For quality control of the genotype data, we removed variants that violated Hardy-Weinberg Equilibrium and had a missing rate of more than 5%, and focused on variants with a minor allele frequency greater than 0.05. From literature review, we found the following known senescence genes: TERT, ATM, CDKN1A, CDKN2B, TP53, IGFBP7, and MAPK10 ^28–37^. In order to identify a more inclusive network of senescence genes using a data-driven approach, we inputted the known senescence genes into GeneMania ^38^, a platform that finds other genes related to those provided, to build a candidate network of senescence genes. GeneMania found a network of 27 genes (appendix p. 9), and 22 of them had evaluable SNPs in our genotype dataset. We ran the Burden test using the GENESIS package ^39^ in R for the gene-based analysis. A gene-based analysis considers the aggregate effect of multiple variants in a test, and we used the burden test to perform this analysis ^40^. The burden test collapses information of variants into a single genetic score ^41^, and the association of this score and the outcome of interest, FaceAge or chronological age, is tested. We provided the effect size with 95% confidence intervals and score test p-values for the genes.

## Data Availability

A part of the data produced in the present study (e.g., data that are not linked to patients) is available upon reasonable request to the authors.

## ACKNOWLEDGEMENTS

The authors acknowledge financial support from NIH (HA: NIH-USA U24CA194354, NIH-USA U01CA190234, NIH-USA U01CA209414, and NIH-USA R35CA22052; BHK: NIH-USA K08DE030216-01), and the European Union – European Research Council (HA: 866504).

## AUTHOR CONTRIBUTIONS

Study conceptualization: O.Z., H.A., R.H.M.; Model design and implementation: O.Z.; Data acquisition, analysis and interpretation: O.Z., D.Bon., H.A., D. Bitt., R.H.M., D.R.; A.D. Writing of the manuscript: O.Z., D.Bon., H.A., R.H.M.; Critical revision of the manuscript: All authors; Statistical Analyses: O.Z., D.Bon., H.A.; Study supervision: H.A., R.H.M.

## ETHICAL COMPLIANCE

This study adheres to ethical principles for human research outlined in the Declaration of Helsinki, and the study and its protocols were approved by the independent hospital ethics review boards at Mass General Brigham (MGB) and Dana-Farber Harvard Cancer Center, Boston MA, and Maastricht University, Maastricht, The Netherlands. All clinical data were handled in compliance with respective institutional research policies.

## Supplementary Appendix

### Supplementary Information 1

Datasets

### Discovery Datasets

For training we used the IMDb-Wiki database <u>(*42*)</u>, a publicly available age-labeled online database of 523,051 face images altogether. An independently curated database of age-labeled face images (that also contained gender and ethnicity labels), the UTKFace (UTK) database <u>(*18*)</u>, was used to evaluate the technical performance of the model. The UTK database contains 24,109 images in total, subdivided into three datasets. Together, these training and testing databases contain photos of known individuals (in particular politicians, actors, professional athletes and other well-known people) in addition to photos of other people in the public domain whose birth dates can be verified. All photographs are labeled with the photo date and birthdate of the individual so that the age at the moment of the photograph can be determined. For the training dataset, 56,304 images from the reference IMDB-Wiki database were selected after applying exclusion criteria, using randomization and augmentation with rebalancing, and performing manual quality assurance on images with age labels of 60 years or older. No clinical patient datasets were used in model training. We randomly rebalanced the training dataset with augmentation using coordinate deformation, horizontal flips, and up to 20 degrees rotation either way, to create a uniformly distributed training set over the age range of 18 to 105, targeting a per-age-year sample size of between 600-700 images. As the dataset was too large to perform manual quality assurance on all images, manual curation and image quality assurance was performed on the training images with age labels of 60 or older because that age group is the most relevant clinically with respect to the oncology datasets we tested (comprising of ∼15,000 of the training images), and to ensure the model would perform at its best over in this age range. In terms of criteria for manual quality assurance, we removed images that were of poor resolution, had artifacts or distortions, or in which the face was covered either completely or partially, or in which there was no face present. For technical validation we assessed the performance of FaceAge across genders and ethnicities in the presumed healthy individuals included in the UTK dataset. After manual quality assurance and curation, data of 2,547 individuals were included in subsequent analyses. Only age-labeled photos of real people were used in model training and validation; the website https://thispersondoesnotexist.com served to generate example face photos for illustrative purposes and figure creation, so as to not publish face photos of real people, but was not used in any technical capacity.

### Clinical Datasets

Three large retrospective oncology datasets from separate institutions were used for testing of the FaceAge algorithm totaling 6,196 cancer patients in the final analysis. Two smaller datasets of non-cancerous patients totaling 535 patients were used as a control for validation purposes. Cancer patients were allowed to have had multiple courses of radiotherapy, as well as surgery and/or systemic therapy, although curatively treated patients had only a single course of radiation treatment. All face photographs used for the analyses of patients treated curatively were acquired prior to the patient’s first treatment. Patients were excluded if no treatment registration photographs were available or of poor-quality, or if their registration date, treatment date, or photo date did not correspond within three months of each other.

#### MAASTRO Cohort

The first clinical dataset consists of 6,835 patients with a cancer diagnosis of which data was prospectively collected and included in the MAASTRO Biobank (Maastricht, The Netherlands). These patients were treated with both curative and palliative intent between 2006 and 2019. The predominant primary malignancies amongst these patients were breast, colorectal, prostate, lung and head and neck cancer. After eliminating records of patients with missing face images, duplicate records, records of patients without follow-up information, and manual image quality assessment, a total of 5,498 entries remained. After removing records of metastatic and/or patients treated with palliative intent or for ductal carcinoma in-situ of the breast (DCIS), the final cohort contained data of 4,906 patients.

#### Harvard Thoracic Cohort

The second clinical dataset consists of 2035 records of thoracic cancer patients who had their most recent treatment with radiotherapy at Dana Farber – Brigham and Women’s Cancer Center between 2008 and 2018. The predominant histology was adenocarcinoma (a form of non-small cell lung cancer) and most patients had Stage III cancer (based on AJCC 7^th^ edition). After eliminating duplicate records and applying exclusion criteria, 802 records remained. Manual image quality assurance and curation reduced the number to a final analysis cohort of 573 patients.

#### Harvard Palliative Cohort

The third clinical dataset consists of 1775 records of palliative patients with metastatic disease seen for consideration of palliative-intent treatment at Dana Farber – Brigham and Women’s Cancer Center between 2008 and 2020. The predominant primary malignancies amongst these palliative patients were lung, breast, prostate and colorectal cancer. After removing duplicate records, records of patients who ended up not receiving treatment, and records with inconsistent dates and/or missing or poor-quality face images, 717 patients remained for subsequent analyses.

#### Harvard Non-cancerous Cohorts

Two smaller cohorts of patients who had their face photographs taken in a clinical setting as part of routine workflow were used as a non-cancerous control to evaluate FaceAge model age predictions, which could then be compared with the predictions from the oncology cohorts. The first cohort consisted of patients treated with benign conditions including, keloids, heterotopic ossification, benign intracranial tumors such as meningiomas and vestibular schwannomas, and cardiovascular conditions, and the second cohort consisted of patients with ductal carcinoma *in situ* of the breast, a precancerous condition that if left untreated leads to development of invasive breast cancer in approximately 30% of patients. The datasets were generated using queries of the electronic medical record systems of Dana Farber – Brigham and Women’s Cancer Center based on clinical indications for radiation therapy, and face photographs were collected between 2009-2023 as part of routine clinical care. The same quality assurance procedure was applied to such datasets before processing (e.g., removal of images with face partially covered with a face mask, error during face extraction phase (MTCNN), etc.), leading to exclusion of 62 and 46 patients for the benign and DCIS cohorts, respectively, resulting in the final inclusion of 112 patients in the benign cohort and 423 patients in the DCIS cohort.

### Supplementary Information 2

Physician Survey

### Physician Survey and Comparison of Human to Machine Performance

A survey was conducted to assess the performance of oncologists and palliative care physicians in estimating the apparent age and 6-month survival of *n* = 100 randomly-selected palliative cancer patients from the Harvard Palliative database, and to compare their performance against FaceAge directly, and to a Cox proportional hazards survival model based on FaceAge. The survey was sent to attending physicians, residents and lay researchers at Harvard-affiliated hospitals. A total of 10 survey participants were enlisted: 5 attending staff, 3 residents and 2 lay researchers. The survey consisted of two parts, administered two weeks apart to reduce memory bias. The first part of the survey presented survey takers with the face photograph of each of the 100 patients, and no accompanying chart information, and the survey-taker then asked to estimate the age of the patient (by decade) and whether the patient would be alive in 6 months’ time <u>(*53*–*55*)</u>. The second part of the survey presented survey-takers with the face photograph accompanied by chart information (without identifiers) that contained the same clinical information available to a Cox PH survival risk model incorporating FaceAge. This risk model was used in the survey to compute a predicted probability of death with respect to time, incorporating the clinical covariates of the TEACHH database, using FaceAge in place of chronologic age, with the same covariates made available to clinicians for survival prediction. The FaceAge risk model was fitted to the remainder of the Harvard Palliative cohort excluding the 100 randomly-selected survey cases, using forward and backward selection of covariates with *p*-value cutoff of 0.2 (see appendix p. 16-17). During the second part of the survey, survey-takers were asked to estimate the probability (in increments of 10%) that the given patient would be alive in 6 months, with all chart information provided. Once their response was given, the FaceAge risk model individualized survival probability curve was then presented to them, and the survey-taker asked to give their estimate of survival probability again, with the survey-taker having the choice of ignoring the new information provided by the FaceAge risk model or modifying their answer accordingly. The area under the receiver operating characteristic curve (AUC) and concordance index (C-index) were used to evaluate and compare the estimates of survey-takers and the FaceAge risk model against ground truth, and groups were compared using the non-parametric two-sided paired Wilcoxon signed rank test. A mock survey case with parts 1 and 2 is presented in the appendix for reference (p. 26). A post-survey questionnaire gathering demographics about the survey-taker, including whether they were an attending or resident, and years of experience, was also included.

### Supplementary Information 3

Results

**Supplementary Figure 1.**
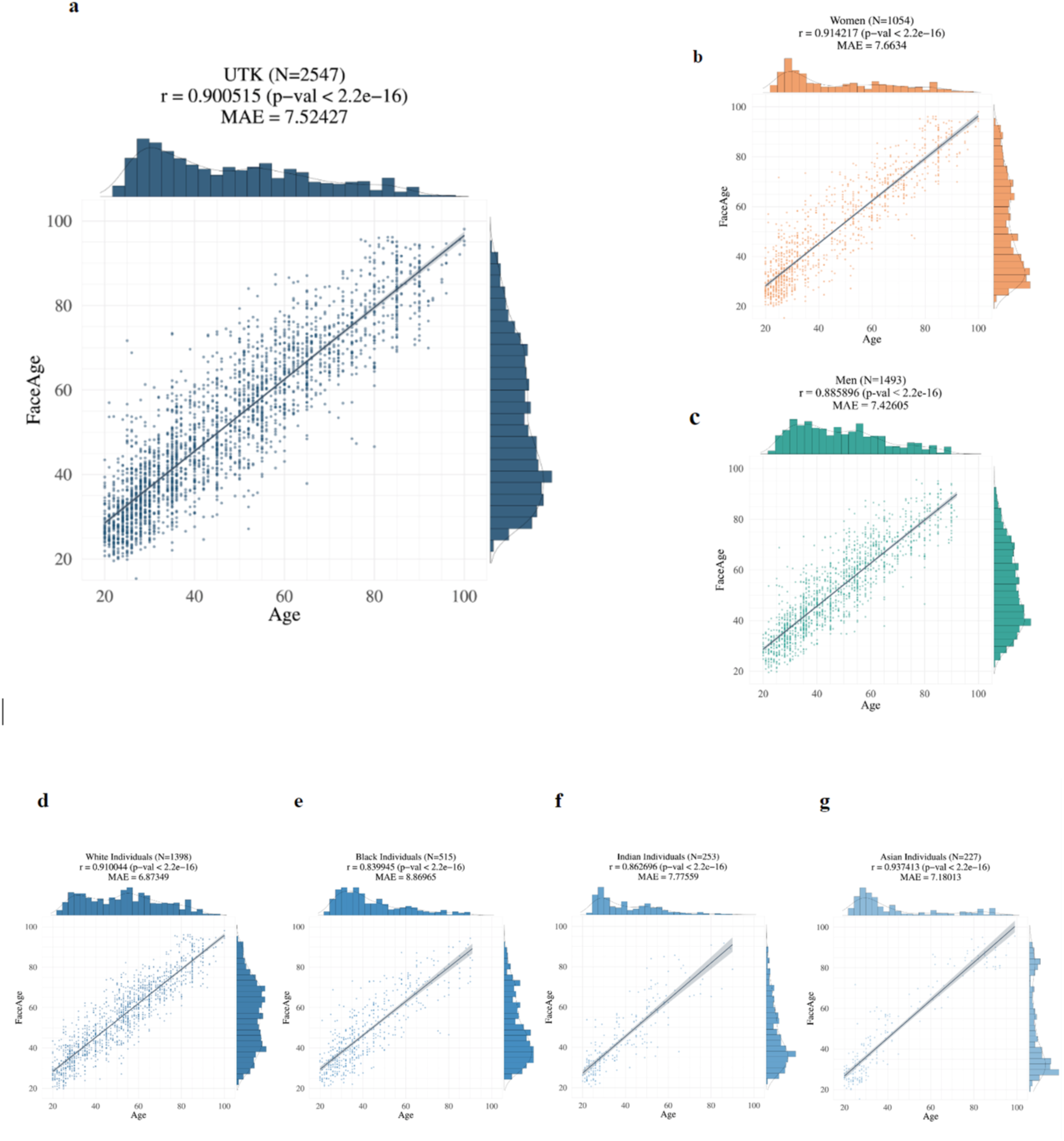
Performance of the FaceAge algorithm in the independent UTK test dataset, for all (**a**), women (**b**), men (**c**), as well as for white (**d**), black (**e**), indian (**f**), and asian (**g**) ethinic subgroups. The model performance is similar and significant across all the groups. *MAE*: Mean Absolute Error, *r*: Pearson R.

**Supplementary Figure 2.**
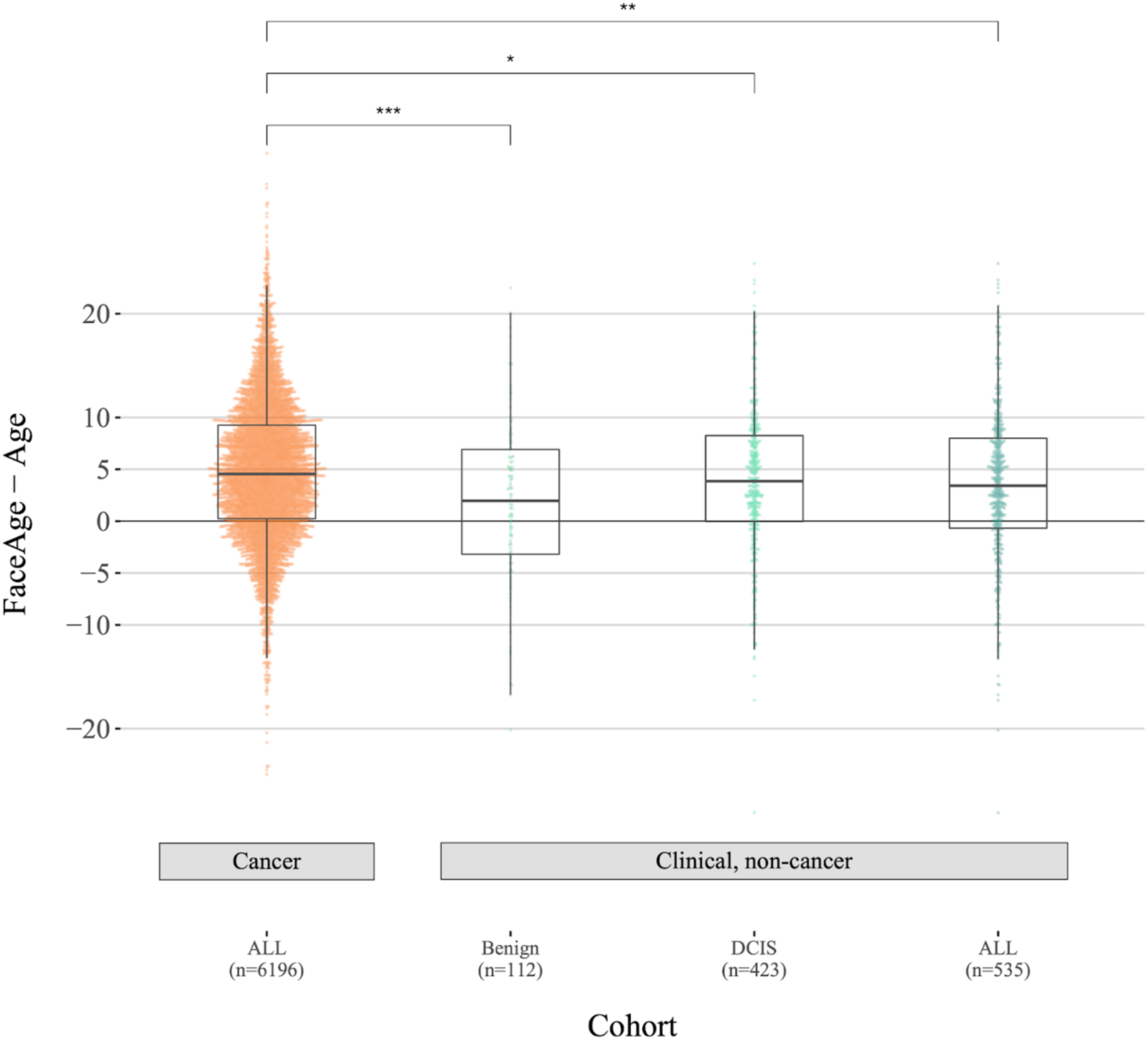
Difference between FaceAge and age in healthy and clinical non-cancer cohorts, to investigate if individuals look older or younger compared to their age. Analyzing all cancer patients included in our analysis, we found that, on average, cancer patients look older than their age (mean 4.79 years, *P* < 0.001). This larger FaceAge-to-chronologic age gap was significant when comparing cancer patients to the reference UTK dataset of healthy individuals 60 years and older (*P* < 0.001) and to the two clinical non-cancer datasets (benign patients: *P* < 0.0001 and DCIS patients: *P* < 0.019) acquired in the same clinical settings and with the same equipment as that of cancer patients, demonstrating that cancer patients look older than those who do not have cancer.

**Supplementary Figure 3.**
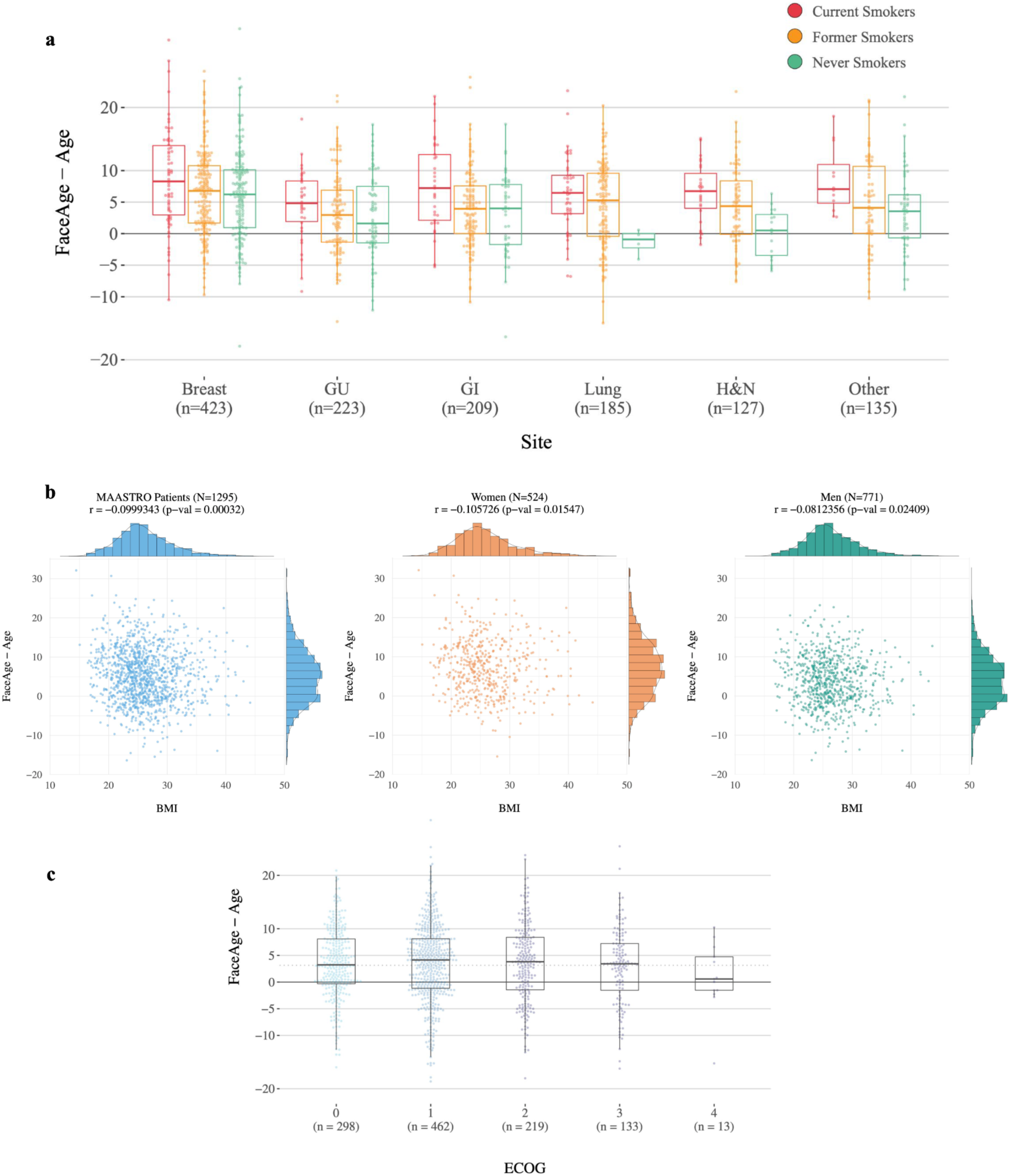
Association of the FaceAge algorithm with lifestyle factors. **a)** Difference between FaceAge and age with the smoking history for different types of cancer patients in the MAASTRO cohort. **b)** Association of the difference between FaceAge and age with body mass index (BMI), for all, women, and men in the MAASTRO cohort. **c)** Association of the difference between FaceAge and age with performance status (Eastern Cooperative Oncology Group (ECOG)) in the HARVARD cohort. We found no significant differences between the groups (unpaired two-sided t-test, P > 0.165). (*r*: pearson R.)

**Supplementary Figure 4.**
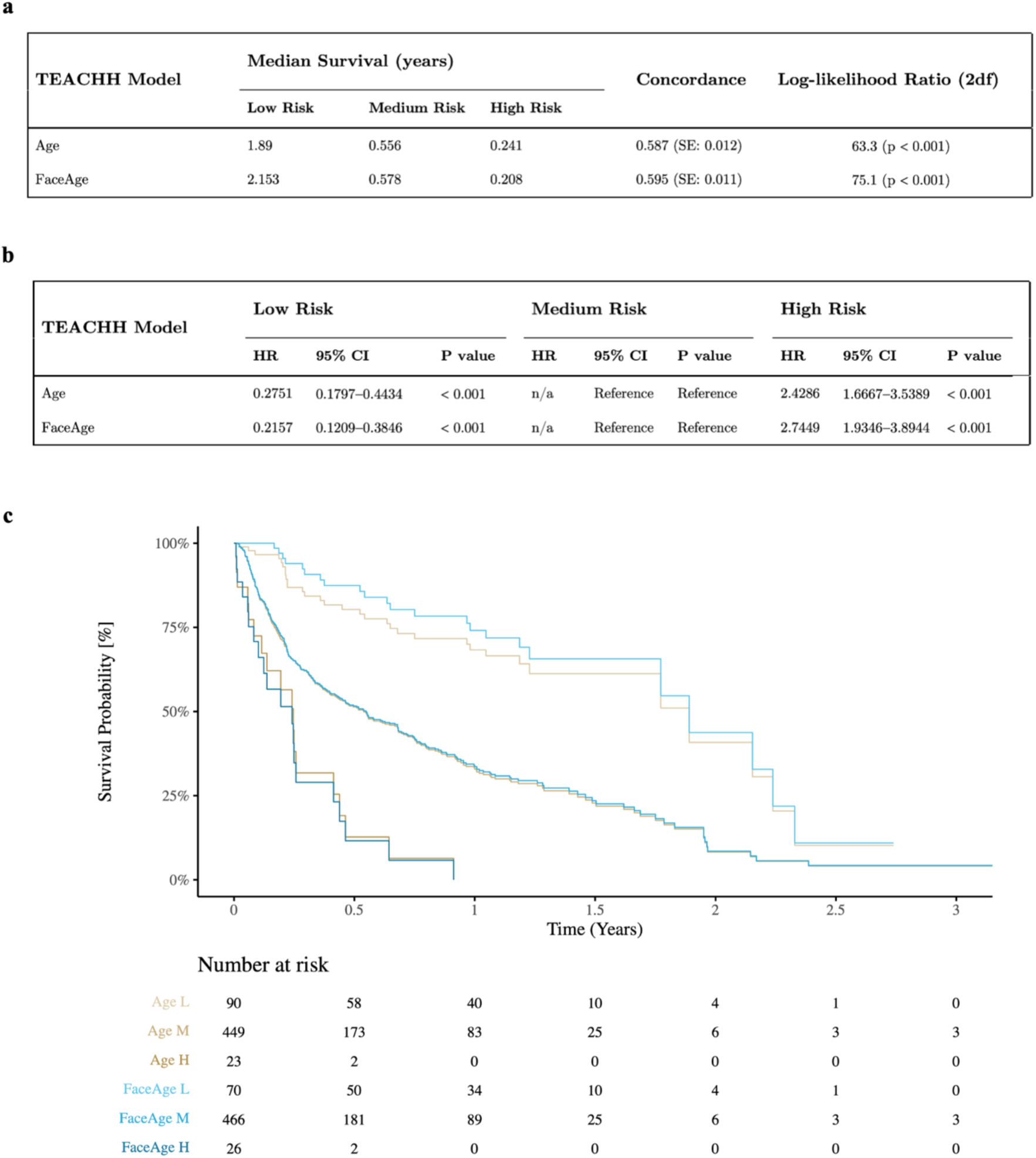
Cox regression analysis of age and FaceAge TEACHH models in the HARVARD palliative cohort. Using FaceAge instead of chronologic age as a covariate in the TEACHH model increases its discriminatory power, as quantified by decreased median survival and increased hazard ratio (HR) of the highest of the risk group, and increased median survival and decreased HR of the lowest risk group. c) TEACHH model Kaplan-Meier survival curves (all-cause mortality) obtained by using chronologic age (solid line) and FaceAge (dashed line) as covariates with 60 years threshold for both. Substituting age with FaceAge, significantly increased the discriminatory power of the model, increasing hazard ratio (HR) of the highest of the risk groups and decreasing the HR of the lowest risk group.

**Supplementary Figure 5.**
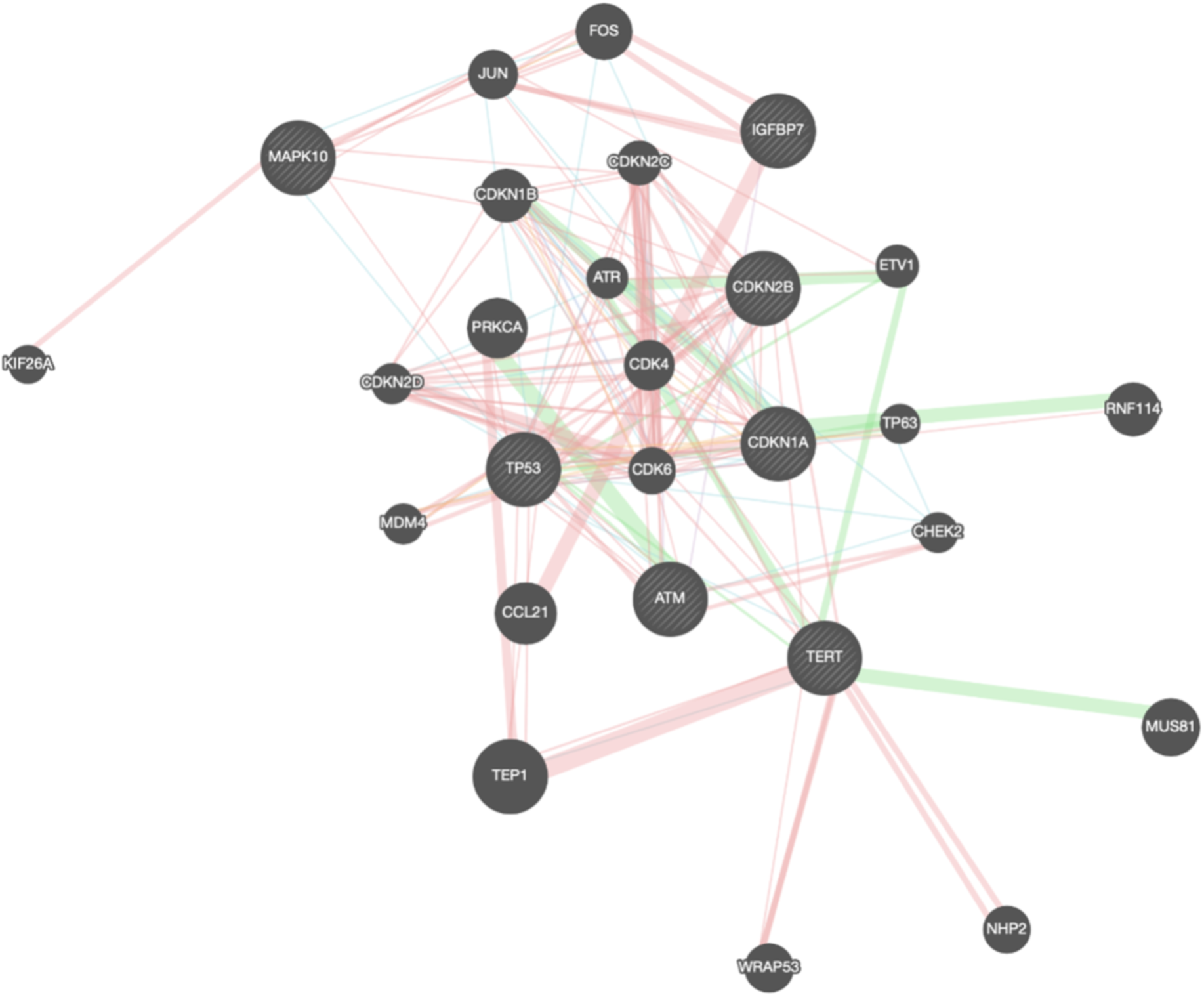
Association of FaceAge and Chronological Age with senescence genes. GeneMania network of senescence genes. Red edges indicate physical interaction, green genetic interactions, and blue pathway. The size of the node represents the score assigned by label propagation algorithms reflecting the strength of association between the node and the input list of genes, i.e., TERT, ATM, CDKN1A, CDKN2B, TP53, IGFBP7, and MAPK10.

**Supplementary Table 1.** Clinical characteristics of the a) MAASTRO Cohort (n=4,906), and b) HARVARD thoracic cohort (N=573). *SBRT*: Stereotactic Body Radioherapy.

| Characteristics |  | Number | Percent |
| --- | --- | --- | --- |
| Sex | Female | 2310 | 47.1 |
| Age at Treatment | Median | 66.0 |  |
|  | Range | 22.0 – 94.0 |  |
| Main Tumor Group | Breast cancer | 1337 | 27.3 |
|  | Gastrointestinal cancer | 1003 | 20.4 |
|  | Genitourinary cancer | 843 | 17.2 |
|  | Lung cancer | 737 | 15.0 |
|  | Head and neck cancer | 456 | 9.3 |
|  | Other types of cancer | 530 | 10.8 |
| Smoking History | Yes (current/former/never) | 1302 | 26.5 |
| BMI | Yes | 1297 | 26.4 |
| ECOG | Yes | 1170 | 23.8 |

**Supplementary Table 1.**
| Characteristics |  | Number | Percent |
| --- | --- | --- | --- |
| Sex | Female | 303 | 52.8 |
| Age at Treatment | Median | 69.0 |  |
|  | Range | 33.3 – 93.2 |  |
| Median Overall Survival (months) | Crude | 14.4 |  |
|  | Actuarial | 16.9 |  |
| Diagnosis | Non-small cell lung cancer | 450 | 78.5 |
|  | Small cell lung cancer | 49 | 10.9 |
|  | Other types of cancer | 74 | 12.9 |
| Treatment Intent | Curative non-SBRT | 433 | 75.6 |
|  | Curative SBRT | 106 | 18.5 |
|  | Palliative | 34 | 5.9 |
| Clinical Stage | I | 145 | 25.3 |
|  | II | 70 | 12.2 |
|  | III | 279 | 48.7 |
|  | IV | 70 | 12.2 |
|  | Not specified | 9 | 1.6 |
| Tumor Grade | ≥ 2 | 322 | 56.2 |
| Race | Caucasian | 493 | 86.0 |
| Smoking History | Yes (current/former/never) | 509 | 88.8 |
| BMI | Yes | 106 | 18.5 |
| ECOG | ≥ 2 | 109 | 19.0 |

**Supplementary Table 2.** Clinical characteristics of the HARVARD palliative cohort (N=717).

| Characteristics |  | Number | Percent |
| --- | --- | --- | --- |
| Sex | Female | 384 | 53.6 |
| Age at Treatment | Median | 65.2 |  |
|  | Range | 19.6 – 97.6 |  |
| Median Overall Survival (months) | Crude | 4.5 |  |
|  | Actuarial | 8.2 |  |
| Time from Diagnosis to Metastasis | Median | 1.0 months |  |
|  | Range | 0 – 26.8 years |  |
| Time from Metastasis to RT Consult | Median | 4.5 months |  |
|  | Range | 0 – 33.0 years |  |
| Primary Diagnosis | Lung | 201 | 28.0 |
|  | Breast | 118 | 16.4 |
|  | Prostate | 58 | 8.1 |
|  | Colorectal | 44 | 6.1 |
|  | Gynecologic | 43 | 6.0 |
|  | Melanoma | 38 | 5.3 |
|  | Esophagus | 24 | 3.3 |
|  | Renal | 24 | 3.3 |
|  | Sarcoma | 24 | 3.3 |
|  | Unknown primary | 20 | 2.8 |
|  | Bladder | 19 | 2.6 |
|  | Head and neck | 10 | 2.6 |
|  | Pancreas | 18 | 2.5 |
|  | Neuroendocrine | 12 | 1.4 |
|  | Cholangiocarcinoma | 9 | 1.3 |
|  | Hepatocellular | 8 | 1.1 |
|  | Non-melanoma skin | 8 | 1.1 |
|  | Stomach | 8 | 1.1 |
|  | GU (non-bladder/testicular) | 6 | 0.8 |
|  | Mesothelioma | 5 | 0.7 |
|  | Testicular | 4 | 0.6 |
|  | Small bowel | 3 | 0.4 |
|  | Other | 6 | 0.8 |
| Metastases | Bone | 344 | 48.0 |
|  | Brain | 265 | 37.0 |
|  | Spine | 256 | 35.7 |
|  | Lung | 223 | 31.1 |
|  | Liver | 222 | 31.0 |
|  | Lymph | 219 | 30.5 |
|  | Adrenal | 65 | 9.1 |
|  | Other | 153 | 21.3 |
| ECOG | 0 – 1 | 309 | 43.1 |
|  | 2 | 135 | 18.8 |
|  | 3 | 110 | 15.3 |
|  | 4 | 11 | 1.5 |
| Prior Palliative RT (courses) | 0 | 627 | 87.5 |
|  | 1 | 72 | 10.0 |
|  | ≥2 | 14 | 2.0 |
| Prior Palliative Chemo (courses) | 0 | 280 | 39.1 |
|  | 1 | 197 | 27.1 |
|  | ≥2 | 237 | 33.1 |
| Hospital Admissions | 0 | 370 | 51.6 |
|  | 1 | 291 | 40.6 |
|  | ≥2 | 55 | 7.7 |
| ER Visits | 0 | 421 | 58.7 |
|  | 1 | 233 | 32.5 |
|  | ≥2 | 62 | 8.7 |

**Supplementary Table 3.**
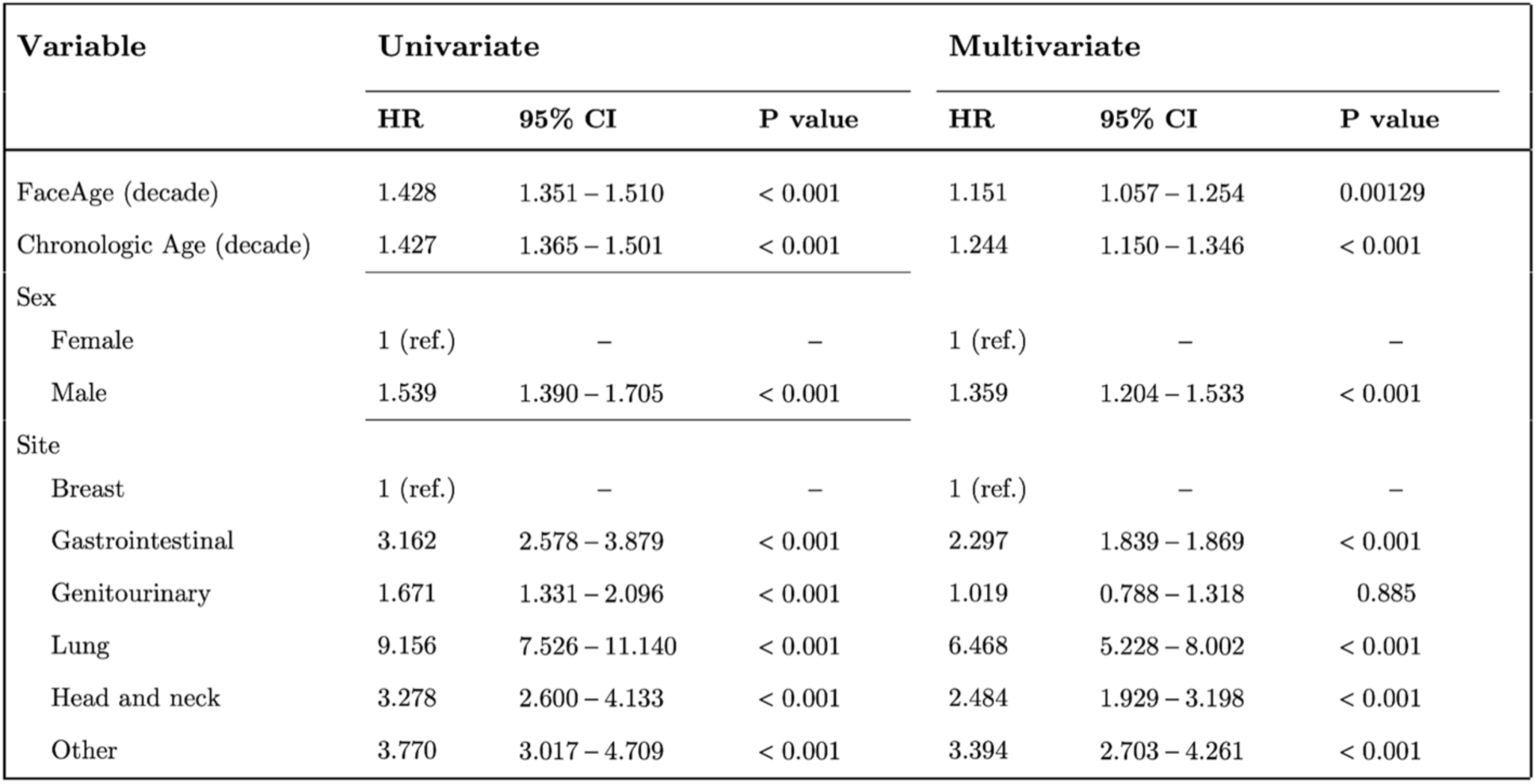
Univariate and multivariate survival analyses of FaceAge on the MAASTRO dataset. FaceAge univariate and multivariate Cox regression analysis for the MAASTRO cohort (N=4,906). Since FaceAge, age, sex, and cancer site have a p-value of less than 0.001 in the univariate analysis, we include all the available covariates in the multivariate model. FaceAge remains statistically significant after adjusting for all the aforementioned covariates. CI: Confidence Interval; *HR*: Hazard Ratio.

**Supplementary Table 4.** Univariate and multivariate survival analyses of FaceAge on the HARVARD Thoracic dataset. FaceAge univariate and multivariate Cox regression analysis for the HARVARD Thoracic cohort (N=573). In the multivariate model, FaceAge remains statistically significant after adjusting for sex, smoking history, ethnicity, treatment intent, tumour grade, ECOG, overall stage, and histology. The same covariates were used to fit a model with age, where age did not have a significant effect. Note, that the multivariate HR for FaceAge in the HARVARD Thoracic dataset is close to the one computed in the multivariate analysis on the MAASTRO cohort (Extended Data Figure 3). BMI was excluded from multivariate analysis as this information was available for only 18.5% of patients. *BMI*: Body Mass Index; *ECOG*: Eastern Cooperative Oncology Group scale; *CI*: Confidence Interval; *HR*: Hazard Ratio.

| Variable | Univariate |  |  | Multivariate (FaceAge) |  |  | Multivariate (Age) |  |  |
| --- | --- | --- | --- | --- | --- | --- | --- | --- | --- |
|  | HR | 95% CI | P value | HR | 95% CI | P value | HR | 95% CI | P value |
| FaceAge (decade) | 1.086 | 0.99 – 1.19 | 0.084 | 1.148 | 1.03 – 1.28 | 0.011 | – | – | – |
| Age (decade) | 1.044 | 0.95 – 1.15 | 0.37 | – | – | – | 1.081 | 0.97 – 1.21 | 0.16 |
| BMI (unit) | 1.00083 | 0.96 – 1.04 | 0.97 | – | – | – | – | – | – |
| Sex |  |  |  |  |  |  |  |  |  |
| Female | 1 (ref.) | – | – | 1 (ref.) | – | – | 1 (ref.) | – | – |
| Male | 1.273 | 1.03 – 1.57 | 0.024 | 1.315 | 1.06 – 1.63 | 0.012 | 1.284 | 1.04 – 1.59 | 0.021 |
| Smoking |  |  |  |  |  |  |  |  |  |
| Never | 1 (ref.) | – | – | 1 (ref.) | – | – | 1 (ref.) | – | – |
| Current/former | 1.261 | 0.84 – 1.87 | 0.26 | 1.014 | 0.67 – 1.54 | 0.95 | 1.064 | 0.70 – 1.62 | 0.77 |
| Ethnicity |  |  |  |  |  |  |  |  |  |
| Caucasian | 1 (ref.) | – | – | 1 (ref.) | – | – | 1 (ref.) | – | – |
| Non-Caucasian | 1.137 | 0.82 – 1.56 | 0.43 | 1.024 | 0.74 – 1.43 | 0.89 | 1.041 | 0.75 – 1.45 | 0.81 |
| Treatment Intent |  |  |  |  |  |  |  |  |  |
| Palliative | 1 (ref.) | – | – | 1 (ref.) | – | – | 1 (ref.) | – | – |
| Curative | 0.458 | 0.3 – 0.69 | < 0.001 | 0.538 | 0.34 – 0.86 | 0.0096 | 0.554 | 0.35 – 0.89 | 0.014 |
| Tumor Grade |  |  |  |  |  |  |  |  |  |
| 1 | 1 (ref.) | – | – | 1 (ref.) | – | – | 1 (ref.) | – | – |
| > 1 | 1.157 | 0.94 – 1.43 | 0.18 | 1.086 | 0.86 – 1.37 | 0.49 | 1.069 | 0.85 – 1.35 | 0.58 |
| ECOG |  |  |  |  |  |  |  |  |  |
| ≤ 1 | 1 (ref.) | – | – | 1 (ref.) | – | – | 1 (ref.) | – | – |
| > 1 | 1.295 | 1.01 – 1.66 | 0.039 | 1.340 | 1.02 – 1.76 | 0.034 | 1.375 | 1.05 – 1.80 | 0.021 |
| Overall Stage |  |  |  |  |  |  |  |  |  |
| I | 1 (ref.) | – | – | 1 (ref.) | – | – | 1 (ref.) | – | – |
| II | 0.822 | 0.59 – 1.14 | 0.24 | 1.179 | 0.79 – 1.75 | 0.42 | 1.129 | 0.76 – 1.68 | 0.55 |
| III | 1.293 | 1.05 – 1.59 | 0.016 | 1.667 | 1.25 – 2.23 | < 0.001 | 1.605 | 1.20 – 2.15 | 0.0015 |
| IV | 1.542 | 1.12 – 2.12 | 0.0074 | 2.029 | 1.33 – 3.09 | 0.0010 | 2.000 | 1.30 – 3.05 | 0.0016 |
| Histology |  |  |  |  |  |  |  |  |  |
| Other | 1 (ref.) | – | – | 1 (ref.) | – | – | 1 (ref.) | – | – |
| NSCLC | 1.053 | 0.84 – 1.32 | 0.66 | 1.221 | 0.92 – 1.62 | 0.16 | 1.222 | 0.92 – 1.62 | 0.16 |
| SCLC | 1.577 | 1.12 – 2.21 | 0.009 | 1.785 | 1.16 – 2.74 | 0.0083 | 1.781 | 1.16 – 2.74 | 0.0085 |

**Supplementary Table 5.** Univariate and multivariate survival analyses for FaceAge and Age on the HARVARD Palliative dataset. FaceAge univariate and multivariate Cox regression analysis for the HARVARD Palliative cohort (N=717). The covariates for the multivariate model are selected using a forward selection procedure with a p-value cutoff of P < 0.2. In the final multivariate model, FaceAge remains statistically significant after adjusting for sex, smoking history, ethnicity, treatment intent, tumour grade, ECOG, overall stage, and histology, while age was not. Furthermore, the multivariate HR for FaceAge in the HARVARD Palliative dataset is similar to the multivariate analysis on both the MAASTRO and HARVARD Thoracic cohorts (Extended Data Figures 5 and 6). BMI: Body Mass Index; ECOG: Eastern Cooperative Oncology Group scale; CI: Confidence Interval; HR: Hazard Ratio.

| Variable | Univariate |  |  | Multivariate (FaceAge) |  |  | Multivariate (Age) |  |  |
| --- | --- | --- | --- | --- | --- | --- | --- | --- | --- |
|  | HR | 95% CI | P value | HR | 95% CI | P value | HR | 95% CI | P value |
| The multivariate analysis includes all the covariates for which $p < 0.2$ in the univariate analysis | | | | | | | | | |
| FaceAge (decade) | 1.102 | 1.01 – 1.21 | 0.035 | 1.117 | 1.02 – 1.23 | 0.021 | – | – | – |
| Age (decade) | 1.065 | 0.98 – 1.16 | 0.16 | – | – | – | 1.076 | 0.98 – 1.18 | 0.11 |
| Number of ER Admits | 2.323 | 1.86 – 2.90 | < 0.001 | 1.519 | 1.14 – 2.02 | 0.0039 | 1.493 | 1.12 – 1.99 | 0.0058 |
| Number of Hospital Admits | 1.678 | 0.35 – 2.09 | < 0.001 | 1.020 | 0.77 – 1.34 | 0.89 | 1.014 | 0.77 – 1.34 | 0.92 |
| Number of Prior RT | 1.443 | 1.04 – 2.00 | 0.027 | 1.425 | 1.00 – 2.04 | 0.053 | 1.445 | 1.01 – 2.07 | 0.045 |
| Number of Prior Chemo | 1.244 | 0.95 – 1.63 | 0.12 | 1.854 | 1.32 – 2.60 | < 0.001 | 1.910 | 1.36 – 2.68 | < 0.001 |
| Time to Consult (years) | 0.919 | 0.86 – 0.98 | 0.011 | 0.906 | 0.83 – 0.99 | 0.024 | 0.900 | 0.83 – 0.98 | 0.015 |
| Time to 1<sup>st</sup> Metastasis (years) | 0.92 | 0.92 – 0.95 | < 0.001 | 0.944 | 0.91 – 0.98 | 0.0026 | 0.948 | 0.91 – 0.98 | 0.0043 |
| Sex |  |  |  |  |  |  |  |  |  |
| Female | 1 (ref.) | – | – | 1 (ref.) | – | – | 1 (ref.) | – | – |
| Male | 1.021 | 0.82 – 1.27 | 0.85 | – | – | – | – | – | – |
| Ethnicity |  |  |  |  |  |  |  |  |  |
| Caucasian | 1 (ref.) | – | – | 1 (ref.) | – | – | 1 (ref.) | – | – |
| Non-Caucasian | 1.053 | 0.79 – 1.41 | 0.73 | – | – | – | – | – | – |
| ECOG |  |  |  |  |  |  |  |  |  |
| ≤ 1 | 1 (ref.) | – | – | 1 (ref.) | – | – | 1 (ref.) | – | – |
| 2 | 1.852 | 1.45 – 2.37 | < 0.001 | 2.282 | 1.73 – 3.00 | < 0.001 | 2.282 | 1.73 – 3.01 | < 0.001 |
| 3-4 | 2.547 | 1.97 – 3.29 | < 0.001 | 2.762 | 2.03 – 3.75 | < 0.001 | 2.825 | 2.08 – 3.83 | < 0.001 |
| Tumor Site |  |  |  |  |  |  |  |  |  |
| Breast | 1 (ref.) | – | – | 1 (ref.) | – | – | 1 (ref.) | – | – |
| Lung | 1.463 | 1.16 – 1.85 | 0.0016 | 1.690 | 1.98 – 2.65 | 0.022 | 1.783 | 1.14 – 2.28 | 0.011 |
| Prostate | 0.446 | 0.28 – 0.72 | < 0.001 | 1.150 | 0.63 – 2.09 | 0.65 | 1.189 | 0.65 – 2.16 | 0.65 |
| Colorectal | 0.674 | 0.43 – 1.05 | 0.081 | 1.097 | 1.62 – 1.96 | 0.75 | 1.150 | 0.65 – 2.05 | 0.57 |
| Gynecologic | 0.727 | 0.44 – 1.20 | 0.21 | – | – | – | – | – | – |
| Other | 1.574 | 1.15 – 1.78 | < 0.001 | 2.128 | 1.43 – 3.16 | < 0.001 | 2.176 | 1.47 – 3.23 | < 0.001 |
| Bone Metastases |  |  |  |  |  |  |  |  |  |
| No | 1 (ref.) | – | – | 1 (ref.) | – | – | 1 (ref.) | – | – |
| Yes | 1.103 | 0.89 – 1.37 | 0.38 | – | – | – | – | – | – |
| Lung Metastases |  |  |  |  |  |  |  |  |  |
| No | 1 (ref.) | – | – | 1 (ref.) | – | – | 1 (ref.) | – | – |
| Yes | 1.202 | 0.96 – 1.51 | 0.11 | 1.364 | 1.06 – 1.75 | 0.015 | 1.360 | 1.06 – 1.75 | 0.016 |
| Liver Metastases |  |  |  |  |  |  |  |  |  |
| No | 1 (ref.) | – | – | 1 (ref.) | – | – | 1 (ref.) | – | – |
| Yes | 1.321 | 1.06 – 1.65 | 0.015 | 1.458 | 1.15 – 1.86 | 0.0022 | 1.445 | 1.13 – 1.84 | 0.0029 |
| Brain Metastases |  |  |  |  |  |  |  |  |  |
| No | 1 (ref.) | – | – | 1 (ref.) | – | – | 1 (ref.) | – | – |
| Yes | 1.734 | 1.37 – 2.20 | < 0.001 | 1.505 | 1.13 – 2.01 | 0.0054 | 1.508 | 1.13 – 2.01 | 0.0052 |
| Spine Metastases |  |  |  |  |  |  |  |  |  |
| No | 1 (ref.) | – | – | 1 (ref.) | – | – | 1 (ref.) | – | – |
| Yes | 1.277 | 1.02 – 1.59 | 0.03 | 1.189 | 0.93 – 1.52 | 0.17 | 1.190 | 0.93 – 1.52 | 0.17 |
| Lymph Nodes Metastases |  |  |  |  |  |  |  |  |  |
| No | 1 (ref.) | – | – | 1 (ref.) | – | – | 1 (ref.) | – | – |
| Yes | 0.947 | 0.75 – 1.20 | 0.65 | – | – | – | – | – | – |
| Adrenal Metastases |  |  |  |  |  |  |  |  |  |
| No | 1 (ref.) | – | – | 1 (ref.) | – | – | 1 (ref.) | – | – |
| Yes | 2.217 | 1.57 – 3.13 | < 0.001 | 1.495 | 1.02 – 2.18 | 0.037 | 1.466 | 1.01 – 2.14 | 0.047 |
| Other Metastases |  |  |  |  |  |  |  |  |  |
| No | 1 (ref.) | – | – | 1 (ref.) | – | – | 1 (ref.) | – | – |
| Yes | 1.324 | 1.03 – 1.71 | 0.031 | 1.088 | 0.83 – 1.43 | 0.55 | 1.112 | 0.85 – 1.46 | 0.45 |

## REFERENCES

1 Bell CG, Lowe R, Adams PD, et al. DNA methylation aging clocks: challenges and recommendations. Genome Biol 2019; 20: 249.

2 Jones MJ, Goodman SJ, Kobor MS. DNA methylation and healthy human aging. Aging Cell 2015; 14: 924–32.

3 Klutstein M, Nejman D, Greenfield R, Cedar H. DNA Methylation in Cancer and Aging. Cancer Res 2016; 76: 3446–50.

4 Aubert G, Lansdorp PM. Telomeres and Aging. Physiological Reviews 2008; 88: 557–79.

5 Blackburn EH, Greider CW, Szostak JW. Telomeres and telomerase: the path from maize, Tetrahymena and yeast to human cancer and aging. Nat Med 2006; 12: 1133–8.

6 Levy MZ, Allsopp RC, Futcher AB, Greider CW, Harley CB. Telomere end-replication problem and cell aging. Journal of Molecular Biology 1992; 225: 951–60.

7 Weindruch R, Kayo T, Lee C-K, Prolla TA. Gene expression profiling of aging using DNA microarrays. Mechanisms of Ageing and Development 2002; 123: 177–93.

8 McCarroll SA, Murphy CT, Zou S, et al. Comparing genomic expression patterns across species identifies shared transcriptional profile in aging. Nat Genet 2004; 36: 197–204.

9 Karasik D, Hannan MT, Cupples LA, Felson DT, Kiel DP. Genetic Contribution to Biological Aging: The Framingham Study. The Journals of Gerontology: Series A 2004; 59: B218–26.

10 Jain R, Duval S, Adabag S. How Accurate Is the Eyeball Test? Circulation: Cardiovascular Quality and Outcomes 2014; 7: 151– 6.

11 Glare P, Virik K, Jones M, et al. A systematic review of physicians’ survival predictions in terminally ill cancer patients. BMJ 2003; 327: 195.

12 Cheon S, Agarwal A, Popovic M, et al. The accuracy of clinicians’ predictions of survival in advanced cancer: a review. Annals of Palliative Medicine 2016; 5: 9.

13 White N, Reid F, Harris A, Harries P, Stone P. A Systematic Review of Predictions of Survival in Palliative Care: How Accurate Are Clinicians and Who Are the Experts? PLoS One 2016; 11: e0161407.

14 Xia X, Chen X, Wu G, et al. Three-dimensional facial-image analysis to predict heterogeneity of the human ageing rate and the impact of lifestyle. Nat Metab 2020; 2: 946–57.

15 Antony A, Dhandharia S, Gupta S, Azhagiri M. Application Of Deep Learning In Analysis Of Age Groups Based On Social Media Interactions. 2019; 8.

16 Nguyen T, Phung D, Adams B, Venkatesh S. Prediction of Age, Sentiment, and Connectivity from Social Media Text. In: Bouguettaya A, Hauswirth M, Liu L, eds. Web Information System Engineering – WISE 2011. Berlin, Heidelberg: Springer, 2011: 227–40.

17 Kuang L, Pobbathi S, Mansury Y, Shapiro MA, Gurbani VK. Predicting age and gender from network telemetry: Implications for privacy and impact on policy. PLOS ONE 2022; 17: e0271714.

18 Zhang K, Zhang Z, Li Z, Qiao Y. Joint Face Detection and Alignment Using Multitask Cascaded Convolutional Networks. IEEE Signal Processing Letters 2016; 23: 1499–503.

19 Szegedy C, Ioffe S, Vanhoucke V, Alemi AA. Inception-v4, Inception-ResNet and the Impact of Residual Connections on Learning. In: Thirty-First AAAI Conference on Artificial Intelligence. 2017. https://www.aaai.org/ocs/index.php/AAAI/AAAI17/paper/view/14806 (accessed Oct 2, 2021).

20 LFW: Results. http://vis-www.cs.umass.edu/lfw/results.html (accessed Oct 2, 2021).

21 Sandberg D. Face Recognition using Tensorflow. https://github.com/davidsandberg/facenet/. 2021; published online Oct 2. https://github.com/davidsandberg/facenet (accessed Oct 2, 2021).

22 Bland JM, Altman DG. Survival probabilities (the Kaplan-Meier method). BMJ 1998; 317: 1572–80.

23 Bland JM, Altman DG. The logrank test. BMJ 2004; 328: 1073.

24 Collett D. Modelling Survival Data in Medical Research, 3rd edn. CRC Press, 2015.

25 Heagerty PJ, Zheng Y. Survival Model Predictive Accuracy and ROC Curves. Biometrics 2005; 61: 92–105.

26 Hajian-Tilaki K. Receiver Operating Characteristic (ROC) Curve Analysis for Medical Diagnostic Test Evaluation. Caspian J Intern Med 2013; 4: 627–35.

27 Krishnan MS, Epstein-Peterson Z, Chen Y-H, et al. Predicting life expectancy in patients with metastatic cancer receiving palliative radiotherapy: The TEACHH model. Cancer 2014; 120: 134–41.

28 van Moorsel CHM. Trade-offs in aging lung diseases: a review on shared but opposite genetic risk variants in idiopathic pulmonary fibrosis, lung cancer and chronic obstructive pulmonary disease. Curr Opin Pulm Med 2018; 24: 309–17.

29 Dressen A, Abbas AR, Cabanski C, et al. Analysis of protein-altering variants in telomerase genes and their association with MUC5B common variant status in patients with idiopathic pulmonary fibrosis: a candidate gene sequencing study. Lancet Respir Med 2018; 6: 603–14.

30 Sclafani A, Worsham CM, McNeill J, Anandaiah A. Advances in Interstitial Lung Disease Genetics. Am J Respir Crit Care Med 2019; 200: 247–9.

31 Newton CA, Oldham JM, Ley B, et al. Telomere length and genetic variant associations with interstitial lung disease progression and survival. Eur Respir J 2019; 53: 1801641.

32 Arimura-Omori M, Kiyohara C, Yanagihara T, et al. Association between Telomere-Related Polymorphisms and the Risk of IPF and COPD as a Precursor Lesion of Lung Cancer: Findings from the Fukuoka Tobacco-Related Lung Disease (FOLD) Registry. Asian Pac J Cancer Prev 2020; 21: 667–73.

33 Bouten RM, Dalgard CL, Soltis AR, Slaven JE, Day RM. Transcriptomic profiling and pathway analysis of cultured human lung microvascular endothelial cells following ionizing radiation exposure. Sci Rep 2021; 11: 24214.

34 Liu X, Shao C, Fu J. Promising Biomarkers of Radiation-Induced Lung Injury: A Review. Biomedicines 2021; 9: 1181.

35 Chen H, Chen H, Liang J, et al. TGF-β1/IL-11/MEK/ERK signaling mediates senescence-associated pulmonary fibrosis in a stress-induced premature senescence model of Bmi-1 deficiency. Exp Mol Med 2020; 52: 130–51.

36 Shamovsky V. PB | HSP90 Chaperone Cycle for Steroid Hormone Receptors (SHR) in the Presence of Ligand. Reactome – a curated knowledgebase of biological pathways. 2017; published online March 27. https://reactome.org/PathwayBrowser/#/R-HSA-3371497&PATH=R-HSA-8953897,R-HSA-2262752 (accessed Feb 4, 2022).

37 AmiGO 2: Gene Product Details for UniProtKB:P53779. http://amigo.geneontology.org/amigo/gene_product/UniProtKB:P53779. 2018; published online July 2. http://amigo.geneontology.org/amigo/gene_product/UniProtKB:P53779 (accessed Feb 4, 2022).

38 Warde-Farley D, Donaldson SL, Comes O, et al. The GeneMANIA prediction server: biological network integration for gene prioritization and predicting gene function. Nucleic Acids Res 2010; 38: W214–220.

39 Gogarten SM, Sofer T, Chen H, et al. Genetic association testing using the GENESIS R/Bioconductor package. Bioinformatics 2019; 35: 5346–8.

40 Chung J, Jun GR, Dupuis J, Farrer LA. Comparison of methods for multivariate gene-based association tests for complex diseases using common variants. Eur J Hum Genet 2019; 27: 811–23.

41 Lee S, Abecasis GR, Boehnke M, Lin X. Rare-Variant Association Analysis: Study Designs and Statistical Tests. Am J Hum Genet 2014; 95: 5–23.

42 UTKFace. UTKFace. https://susanqq.github.io/UTKFace/ (accessed Oct 2, 2021).

43 Krishnan M, Temel JS, Wright AA, Bernacki R, Selvaggi K, Balboni T. Predicting life expectancy in patients with advanced incurable cancer: a review. J Support Oncol 2013; 11: 68–74.

44 Au PC-M, Li H-L, Lee GK-Y, et al. Sarcopenia and mortality in cancer: A meta-analysis. Osteoporos Sarcopenia 2021; 7: S28–33.

45 Al-Sawaf O, Weiss J, Skrzypski M, et al. Body composition and lung cancer-associated cachexia in TRACERx. Nat Med 2023; 29: 846–58.

46 Christakis NA, Iwashyna TJ. Attitude and Self-reported Practice Regarding Prognostication in a National Sample of Internists. Archives of Internal Medicine 1998; 158: 2389–95.

47 Christakis NA, Smith JL, Parkes CM, Lamont EB. Extent and determinants of error in doctors’ prognoses in terminally ill patients: prospective cohort study Commentary: Why do doctors overestimate? Commentary: Prognoses should be based on proved indices not intuition. BMJ 2000; 320: 469–73.

48 Lamont EB, Christakis NA. Prognostic Disclosure to Patients with Cancer near the End of Life. Ann Intern Med 2001; 134: 1096–105.

49 Garcia RV, Wandzik L, Grabner L, Krueger J. The Harms of Demographic Bias in Deep Face Recognition Research. In: 2019 International Conference on Biometrics (ICB). 2019: 1–6.

50 Kärkkäinen K, Joo J. FairFace: Face Attribute Dataset for Balanced Race, Gender, and Age for Bias Measurement and Mitigation. In: 2021 IEEE Winter Conference on Applications of Computer Vision (WACV). 2021: 1547–57.

51 Bacchini F, Lorusso L. Race, again: how face recognition technology reinforces racial discrimination. *Journal of Information*, Communication and Ethics in Society 2019; 17: 321–35.

52 Pavanello S, Campisi M, Fabozzo A, et al. The biological age of the heart is consistently younger than chronological age. Sci Rep 2020; 10: 10752.

53 Lehallier B, Gate D, Schaum N, et al. Undulating changes in human plasma proteome profiles across the lifespan. Nat Med 2019; 25: 1843–50.

54 Negasheva MA, Zimina SN, Lapshina NE, Sineva IM. Express Estimation of the Biological Age by the Parameters of Body Composition in Men and Women over 50 Years. Bull Exp Biol Med 2017; 163: 405–8.

55 Bana B, Cabreiro F. The Microbiome and Aging. Annual Review of Genetics 2019; 53: 239–61.

56 Lu MT, Ivanov A, Mayrhofer T, Hosny A, Aerts HJWL, Hoffmann U. Deep Learning to Assess Long-term Mortality From Chest Radiographs. JAMA Network Open 2019; 2: e197416.

57 Orlov NV, Makrogiannis S, Ferrucci L, Goldberg IG. Differential Aging Signals in Abdominal CT Scans. Academic Radiology 2017; 24: 1535–43.

58 Cole JH, Poudel RPK, Tsagkrasoulis D, et al. Predicting brain age with deep learning from raw imaging data results in a reliable and heritable biomarker. NeuroImage 2017; 163: 115–24.

59 IMDB-WIKI – 500k+ face images with age and gender labels. https://data.vision.ee.ethz.ch/cvl/rrothe/imdb-wiki/ (accessed Oct 2, 2021).

60 van Lummel EV, Ietswaard L, Zuithoff NP, Tjan DH, van Delden JJ. The utility of the surprise question: A useful tool for identifying patients nearing the last phase of life? A systematic review and meta-analysis. Palliat Med 2022; 36: 1023–46.

61 Verhoef M-J, de Nijs EJM, Fiocco M, Heringhaus C, Horeweg N, van der Linden YM. Surprise Question and Performance Status Indicate Urgency of Palliative Care Needs in Patients with Advanced Cancer at the Emergency Department: An Observational Cohort Study. Journal of Palliative Medicine 2020; 23: 801–8.

62 Hadique S, Culp S, Sangani RG, et al. Derivation and Validation of a Prognostic Model to Predict 6-Month Mortality in an Intensive Care Unit Population. Annals ATS 2017; 14: 1556–61.

